# An economic evaluation of routine hepatocellular carcinoma surveillance for high-risk patients using a novel approach to modelling competing risks

**DOI:** 10.1101/2024.02.20.24303111

**Authors:** Joachim Worthington, Emily He, Michael Caruana, Stephen Wade, Barbara de Graaff, Anh Le Tuan Nguyen, Jacob George, Karen Canfell, Eleonora Feletto

**Affiliations:** The Daffodil Centre, The University of Sydney, a joint venture with Cancer Council NSW; Storr Liver Centre, The Westmead Institute for Medical Research, Westmead Hospital and University of Sydney; Menzies Institute for Medical Research, The University of Tasmania, Hobart, Tasmania; WHO Collaborating Centre for Viral Hepatitis, The Peter Doherty Institute for Infection and Immunity

## Abstract

**Introduction:** Liver cancer is the only cancer in Australia for which mortality rates have consistently risen, despite tests to identify high-risk individuals. Hepatocellular carcinoma (HCC) is the most common form of primary liver cancer. Curative treatment for HCC is typically only available if detected early. Australian clinical guidelines recommend routine 6-monthly ultrasound surveillance, with or without serum alpha-fetoprotein, for individuals with liver cirrhosis. This study assesses the health and economic implications of this recommendation, utilizing novel modeling techniques.

**Methods:** We designed the *sojourn time density model* mathematical framework to develop a model of the evolving risk of HCC, liver disease, and death based on time since diagnosis, incorporating data on liver decompensation, HCC incidence, and HCC survival, and the impact of surveillance on cancer stage and survival.

**Results:** We estimated that adherence to 6-monthly ultrasound, with or without alpha-fetoprotein, can increase early-stage diagnosis rates, reducing HCC mortality by 22%. We estimate a cost-effectiveness ratio of $33,850 per quality-adjusted life-year (QALY) saved for 6-monthly ultrasound HCC surveillance, under the $50,000/QALY cost-effectiveness threshold. HCC surveillance was also estimated to be cost-effective at any interval from 3-24 months.

**Conclusions:** These findings support the current clinical guideline recommendation for 6-monthly ultrasound surveillance, affirming its health benefits and cost-effectiveness, and show that alternative surveillance intervals would remain beneficial and cost-effective. Our model may be used to refine surveillance recommendations for other at-risk population subgroups and inform evidence-based clinical practice recommendations, and the framework can be adapted for other epidemiological modelling. Supporting the clinical guidelines and their ongoing development as evidence evolves may be key to reversing increasing HCC mortality rates in Australia, which are predicted to increase by more than 20% by 2040.

## 1 Introduction

### 1.1 Hepatocellular carcinoma and surveillance

Worldwide, liver cancer is the fourth most common cause of cancer death;[1] in Australia, it is the seventh most common cause of cancer-related death[2] driven by increasing trends in both liver cancer incidence and mortality.[3] A key step to improving mortality outcomes is increased detection at early stages, to improve the potential for curative treatment.[4, 5, 6]

Hepatocellular carcinoma (HCC), the most common form of primary liver cancer, can be detected early through routine HCC surveillance. Routine HCC surveillance is typically recommended for high-risk populations.[7] In Australia, clinical guidelines recommend HCC surveillance with six-monthly ultrasound (US) imaging, with the potential addition of alpha-fetoprotein (AFP) blood testing, for people with compensated liver cirrhosis,[8, 9] and some population groups with chronic hepatitis B.

Determining appropriate HCC surveillance recommendations poses significant challenges, including identifying health benefits and long-term costs,[10] appropriate target groups,[11, 12] and optimal technologies and intervals.[13] This is needed to develop an economic case for health system investment.

### 1.2 Modelling and HCC

Epidemiological modelling allows a formal synthesis and extrapolation of data to evaluate the impact of an intervention. This approach has been used to guide investment in cancer control interventions.[14, 15, 16, 17, 18] For HCC, models can synthesise data sources such as time to liver decompensation and HCC in patients with cirrhosis[19, 20] and cirrhosis and HCC survival.[21, 22, 2, 23, 24] This can be combined with clinical data on surveillance outcomes and cost data to provide cost-effectiveness estimates. Economic models of HCC and surveillance are less established than for other cancers,[15] but are crucial for guiding policy and investment in liver cancer control.

Internationally, modelling studies have analysed a range of populations and interventions for liver cancer control; reviews have been published previously.[8, 7, 25] Australian modelling has shown that six-monthly US, with or without AFP, is likely cost-effective for patients with cirrhosis,[10] and biomarker testing can stratify risk to improve cost-effectiveness.[26] Further modelling which captures detailed progression of liver disease, operates at flexible timescales, and incorporates detailed survival data could refine recommendations for surveillance algorithms.

There are a number of unique considerations for modelling HCC and liver disease, such as the interaction of competing risks of liver decompensation and other liver disease events,[27, 28] the complex distribution of sojourn times for the development of HCC in cirrhotic patients,[29] significant comorbidities in the target population,[30] and short expected survival times for HCC patients.[31] Simple Markov-style models struggle to capture this complexity, while more complex models such as discrete event simulations[32, 33, 34], agent-based/microsimulation models,[35, 14] and semi-Markov models[36, 37, 38] can be hampered by a lack of data, and typically introduce stochastic elements and increase computation burden. In the absence of detailed health state data, using observable data on the time an individual takes to transition between health states, the *sojourn time*, can improve model fidelity.

### 1.3 Approach

To address this, we developed the *sojourn time density model* framework. We model the dynamics of the probability density of the sojourn time by health state, tracking both the likelihood of being in a given state and the expected time in that state.

The model is parametrised by the *cause-specific hazard rates* [39, 40] (analogous to *transition intensities*[38]), allowing us to exploit survival data and methods[41] such as Kaplan-Meier estimators[42, 39, 43] and risk ratios.[44] This captures the same detail as typical survival models, with the addition of serial state transitions, and is analogous to semi-Markov and discrete event simulations, but with a deterministic numerical scheme.

### 1.4 Aims

In this manuscript we describe the model structure and numerical scheme for sojourn time density models. We develop a health-economic model of liver disease and HCC, and evaluate the benefits and costs routine HCC surveillance through six-monthly liver US, with or without AFP testing, compared to no surveillance. We also provide an analysis of the potential impact of varying surveillance intervals.

The modelling described in this manuscript was developed to inform the *Clinical practice guidelines for HCC surveillance for people at high risk in Australia*.[8] The development of these guidelines was led by clinicians in the area of liver disease and liver cancer alongside a multi-disciplinary working party, including healthcare and clinical representatives, representatives with lived experience and other community representatives. This group also informed the aims and structure of this economic analysis.

## 2 Methods: Sojourn Time Density Model

We now describe the sojourn time density model framework and demonstrate its useful properties. We also develop a numerical scheme for the calculation.

### 2.1 Sojourn time density modelling

Consider a compartmental model with *N* possible discrete states labelled 1, 2, …, *N*, such that at any time *t ∈T* (typically *T* = [0, *∞*)) an individual belongs to one of these states. We describe not only the likelihood of an individual being in a given state, but also track the distribution of the length of time spent in that state, the *sojourn time τ ∈ T*.

Doing so describes a continuous-time random process *{X*_*t*_*}*_*t∈T*_ with discrete state space *{*1, 2, …, *N }*. This process is defined such that *P* (*X*_*t*_ = *i*) = *P* (individual in state *i* at time *t*). We can equivalently define the *jump process {Y*_*n*_, *T*_*n*_*}*_*n*=0,1,2,…_, such that *Y*_*n*_ = *X*_*t*_ *∀ t ∈* [*T*_*n*_, *T*_*n*+1_), *Y*_*n*_ ≠ *Y*_*n*+1_.[45]

The *cause-specific hazard rates*

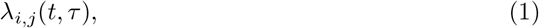

depending on both the model time *t* and sojourn time *τ*, are the key parameters for the model. These are the instantaneous transition rates for an individual in state *i* to transition to state *j*, at time *t*, given that the individual has spent time *τ* in *i* without any transitions (i.e, entered state *i* at time *t − τ*). By modelling the distribution of *τ*, we can capture evolving transition rates. The *t* dependency can capture temporal trends as well as evolving risk with age *a* = *a*_0_ + *t* where *a*_0_ is the age at *t* = 0. If there is no possible transition between *i* and *j*, then *λ*_*i,j*_ = 0.

Formally, the following should hold:

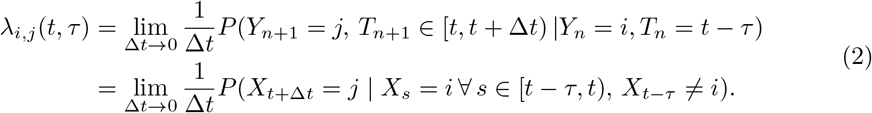

Compare this to the equivalent definition for semi-Markov models, Equation (3) in Krol et al, 2015[46]. For convenience assume *λ*_*i,i*_(*t, τ*) = 0 (i.e. there are no self-loops).

To track the model state and the sojourn time, we introduce the *density functions for the sojourn time*

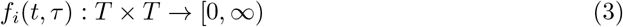

for *i* = 1, 2, …, *N*. These are the density functions for the sojourn time *τ* and the state *i* at time *t*. Explicitly, the set of functions *{f*_*i*_(*t, τ*)*}*_*i*=1,2,…,*N*_ is a *probability density function* on the sample space (*τ, i*) *∈ T ×* {1, 2, …*N* }, describing the likelihood of being in a particular state with a particular sojourn time at fixed time *t ∈ T*.

Then we can define the probability mass of *X*_*t*_

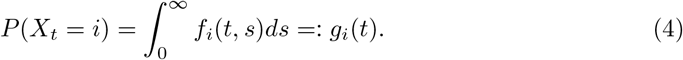

We should expect that

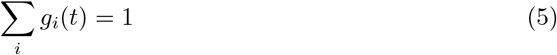

i.e. the total mass probability is 1.

Our goal now is to describe the dynamics of *f*_*i*_(*t, τ*), and then show this definition satisfies (2) and (5). Let *f*_*i*_(*t, τ*) satisfy the partial differential equation

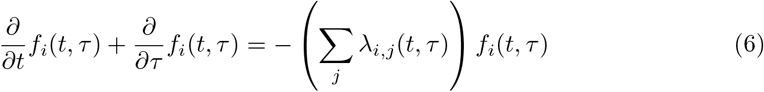

with boundary condition

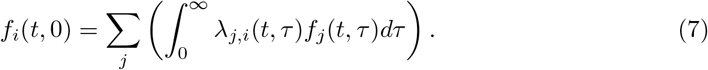

The LHS of equation (6) is motivated by the transport equation along lines of constant *t − τ*, representing the likelihood of remaining in state *i* while both *t* and *τ* increase, while the RHS captures the likelihood of making a transition to a subsequent state. This can be seen by observing that along the directional derivative

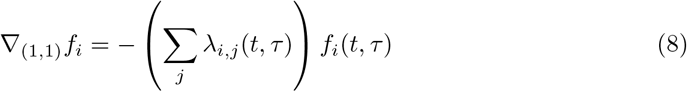

the likelihood decays proportionally to the hazard rate (see Section 2.2). The boundary condition (7) represents the accumulation of probability mass in state *i* post-transition. By (5), the initial conditions *f*_*i*_(0, *τ*) should satisfy

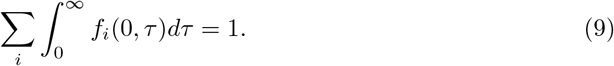

Additionally,

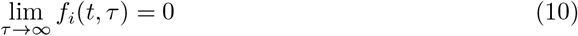

(i.e. the model is supported on finite time only; see Section 2.4). For a further discussion of initial conditions, see Appendix C.1.

We now have a complete description of the random process *X*_*t*_ through (4), (6), (7), and appropriate initial conditions. This constitutes what we will call a *sojourn time density model*. These models resemble semi-Markov processes/Markov renewal processes,[38] as the sequence of transitions is Markov (as the next state depends only on the previous state and transition time) but the time between transitions is not.

We now show that the process defined by (4) is well-defined and driven by the hazards *λ*_*i,j*_(*t, τ*) as expected.

**Theorem 2.1**.

*The random process defined by* (4) *satisfies conditions* (2) *and* (5). *Proof*. We calculate the derivative of *g*_*i*_(*t*), i.e. the rate of change in likelihood of being in state *i* at time *t*

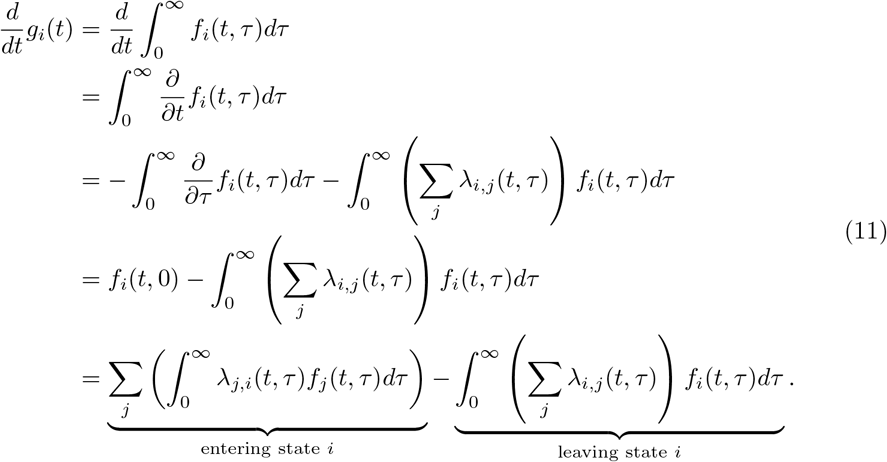

This satisfies the definition the hazard rate - the likelihood of being in state *i* decreases proportionally to the hazard of every transition leaving state 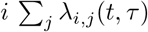, and increases for every transition entering state 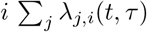 proportional to the weight of those states *f*_*j*_(*t, τ*).

Note that

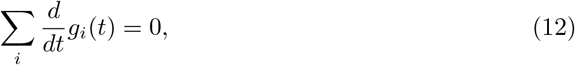

i.e. that the total probability mass is conserved. Therefore with initial conditions satisfying equation (9), (5) is satisfied.

If we know that the system enters state *i* at time *t*^*∗*^, i.e. (*Y*_*n*_, *T*_*n*_) is realised as (*i, t*^*∗*^) and remains there to at least time *t*^*∗*^ + *τ* ^*∗*^ (i.e. *T*_*n*+1_ *− T*_*n*_ *≥ τ* ^*∗*^), this corresponds to

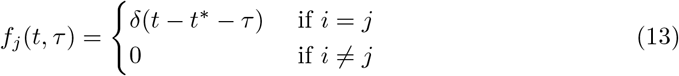

for all *t ∈* [*t*^*∗*^, *t*^*∗*^ + *τ* ^*∗*^) where *δ*(*t*) is the Dirac delta function. Then

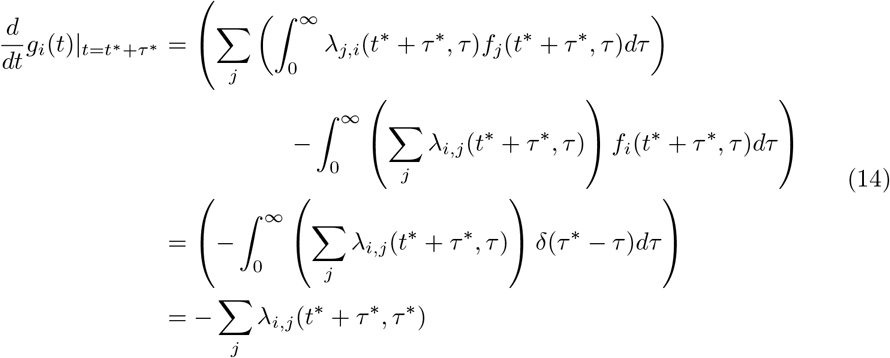

and for *j≠ i*

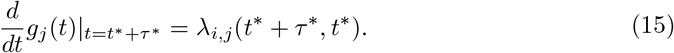

In other words, the likelihood of transition out of state *i* is proportional to the sum of the hazards Σ_*j*_ *λ*_*i,j*_ and the likelihood of transition *i → j* is proportional to *λ*_*i,j*_. Then

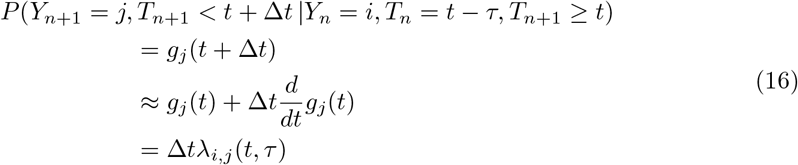

by the above. By taking the limit Δ*t →* 0, (2) is satisfied.

### 2.2 Method of characteristics and survival analysis

We now compute survival curves using method of characteristics. As noted, equation (6) is best interpreted along characteristics of constant *t −τ* corresponding to the state entry time (Figure 1). This allows explicit solutions through method of characteristic, as well as analogies to other survival modelling approaches.^1^

**Figure 1.**
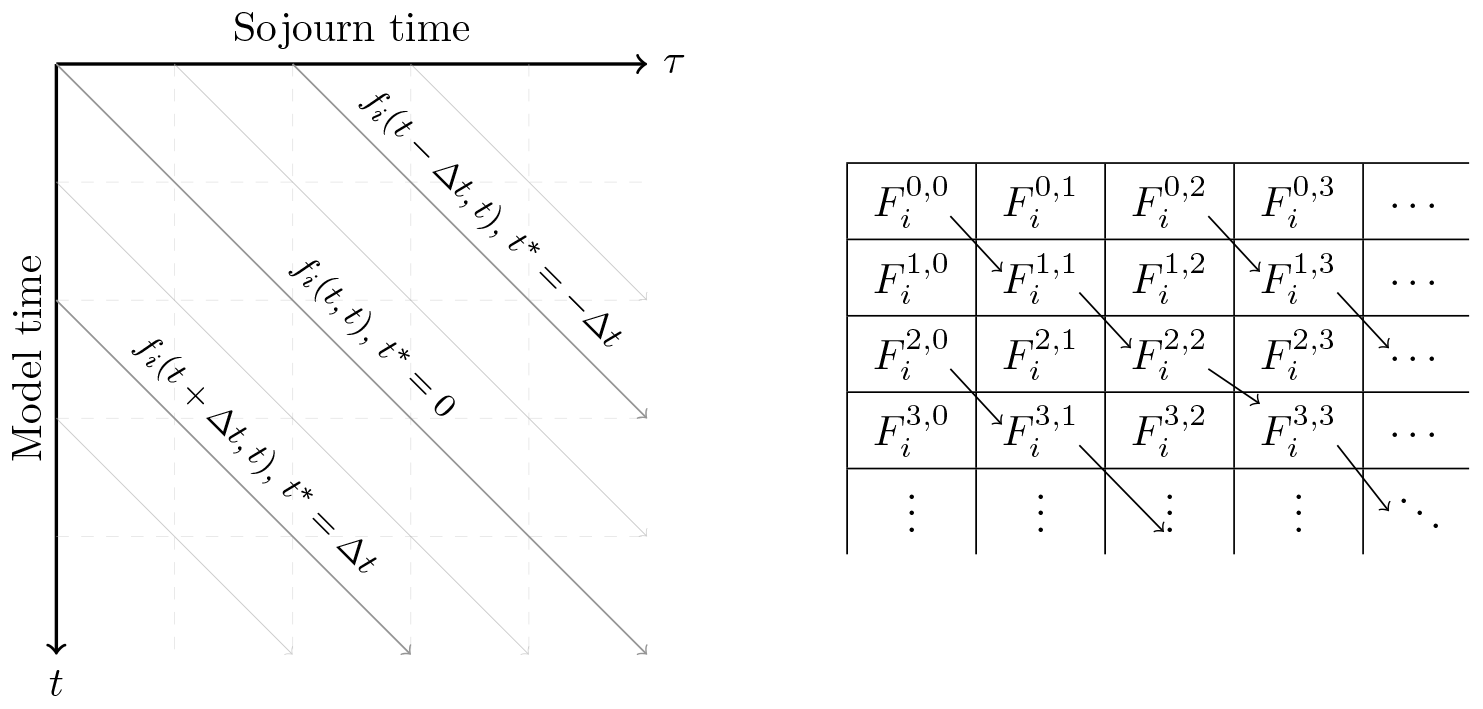
Characteristics of constant t^∗^ = t − τ for f_i_(t, τ) (left) and the corresponding elements 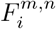 with constant m − n for the numerical scheme (right).

Consider the characteristic *t* = *t*^*∗*^ + *τ* for fixed *t*^*∗*^ *∈* ℝ. Then

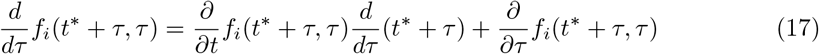

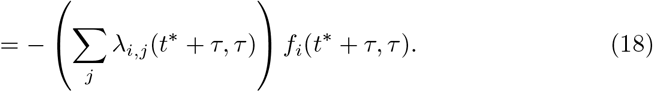

If *t*^*∗*^ *>* 0, this corresponds to a characteristic starting from the boundary; if *t*^*∗*^ ≤ 0, this corresponds to a characteristic from the initial condition. This has solution

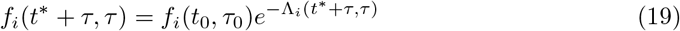

where *t*_0_ = max(*t*^*∗*^, 0), *τ*_0_ = max(*−t*^*∗*^, 0) (so that *t*^*∗*^ = *t*_0_ *− τ*_0_, *t*_0_, *τ*_0_ *≥* 0) and

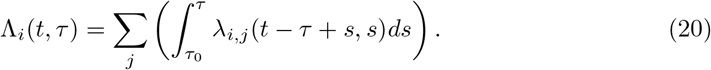

The function Λ_*i*_(*t, τ*) is the *all-cause cumulative hazard* along the characteristic *t −τ* = *t*^*∗*^.Then

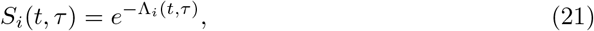

is the *all-cause survival* along this characteristic, and *f*_*i*_(*t*^*∗*^ + *τ, τ*) = *f*_*i*_(*t*_0_, *τ*_0_)*S*_*i*_(*t*^*∗*^ + *τ, τ*). We can correspondingly calculate the *time-to-event distribution* after entering state *i* at time *t − τ* as the distribution

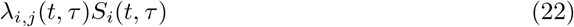

across *j* = 1, 2, …, *N*.

It follows that the probability of the next transition being before time *t* = *t*^*∗*^ + *τ* and also from *i → j*, given entry to state *i* at time *t*_0_, is:

*P* (next transition time *< t ∩* next transition *i → j* | entered state *i* at time *t*_0_)

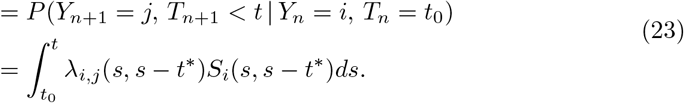

The limit of this is the likelihood of the next transition being to a particular state:

*P* (next transition *i → j* | entered state *i* at time *t*_0_)

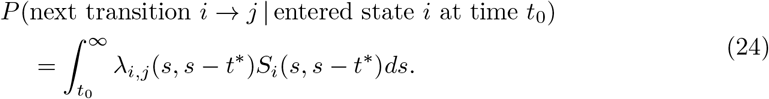

We can also calculate the *cause-specific survival*, assuming other transitions (or events) are censored:

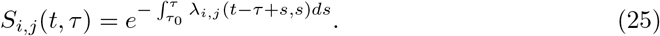

Where cause-specific survival data (equivalently, cause-specific cumulative incidence) is available with competing events censored, this may provide a calibration target for the hazard rates, which can be directly identified from the survival model (see *Hazard and Cumulative hazard* in Clark et al[39]); parametrisation of *λ*_*i,j*_(*t, τ*) across *t*^*∗*^ can then be identified.

Given *t*^*∗*^, the results above are analogous to the functions used for parametric competing risk survival models, time-to-event models, and discrete event models.[47] The sojourn time density model approach generalises this to modelling serial events deterministically. The *all-cause cumulative incidence function* 1 *−S*_*i*_(*t, τ*) is used in e.g. Fine-Gray modelling.[48] A useful guide to translating between cause-specific and all-cause specific hazard rates is included in Asanjarani et al,[38] and shows discrete event simulation distributions can be translated into hazard rates. For discrete event simulations where events do not preclude each other, transitions which preserve sojourn time may be added (Appendix C.4).

A common approach in analytical epidemiological models is to compute likelihood of transition as the convolution of the *τ* -dependent hazard rate and the time of entry.[49, 50] In our formulation, this is reflected in the boundary conditions

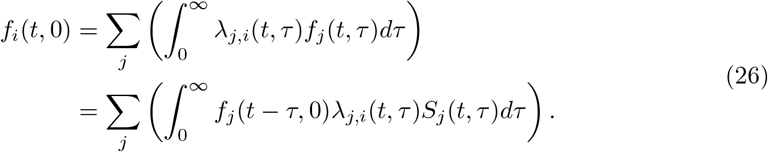

1. If we define entry time distribution *F*_*j*_(*x*) = *f*_*j*_(*x*, 0) and survival-adjusted cause-specific hazard rate *H*(*j, i*)(*x*) = *λ*_*j,i*_(*t, x*)*S*_*j*_(*t, x*), then *F*_*i*_(*t*) = Σ_*j*_ (*F*_*j*_ *∗H*_*j,i*_)(*t*), the convolution of these. We have generalises this approach and incorporated serial transitions.

### 2.3 Numerical scheme

The characteristic solutions developed in Section 2.2 can be used to develop a numerical scheme. For a discretisation of *T* by fixed timesteps Δ*t ∈* ℝ+, define

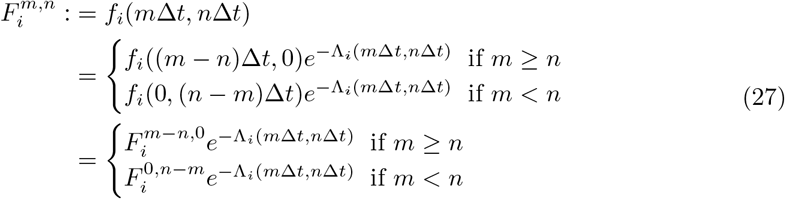

for *m, n ∈* ℤ +. Each value of *m − n* corresponds to a characteristic (*m − n*)Δ*t*. The cumulative hazard functions Λ_*i*_(*t, τ*) may be computed by (20) in simple cases, or estimated iteratively by Δ*t* steps. Equivalently, each element can be calculated iteratively by

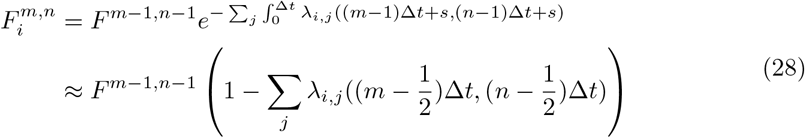

(see Figure 1 for an illustration of this).

For *m < n*, the initial condition is 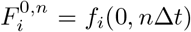 For *m >* 0, the initial condition *F*^*m*,0^ = *f*_*i*_(*m*Δ*t*, 0) is the boundary conditions of the full PDE. A first order scheme can be computed as

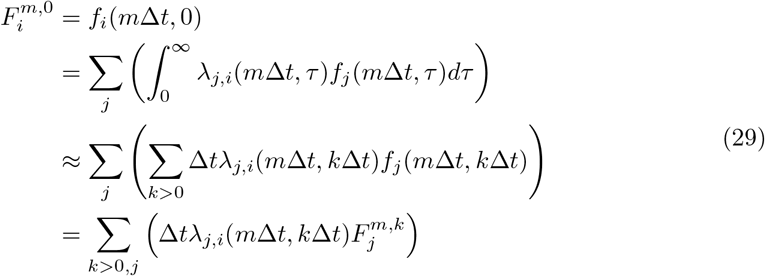

for *m* ≥ 0.

One can subsequently approximate

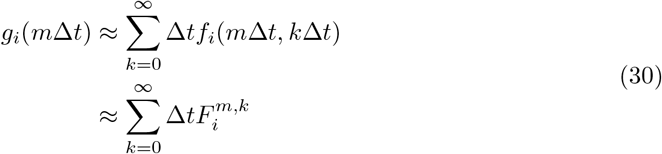

and hence the process *X*_*t*_.

An improvement can be made to this numerical scheme by adjusting the boundary conditions to exactly preserve the total mass at all timesteps:

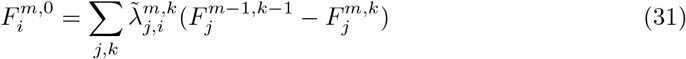

where

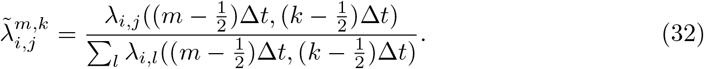

We can then verify that 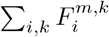 is constant, i.e. the total probability mass Σ_*i*_*g*_*i*_(*t*) is conserved. We have here used the midpoint estimates for *λ*_*i,j*_, which improves accuracy when there is a significant scale difference between hazards.

In epidemiological contexts, hazards are often modelled as continuous and smooth functions, so this first order numerical scheme will have high accuracy.

### 2.4 Hazard functions and common parametric distributions

The sojourn time density modelling scheme is determined by the hazard rates *λ*_*i,j*_(*t, τ*). It is common to describe these rates parameterically.[51] Commonly used parametric forms for these hazard functions include

- constant hazard *λ*_*i,j*_(*t, τ*) = *c*, corresponding to an exponential time-to-event distribution; if all hazards are constant, *X*_*t*_ is Markov.
- polynomial hazard *λ*_*i,j*_(*t, τ*) = *bkτ*^*k−*1^, corresponding to a Weibull distribution, often used to model single-cause mortality.[52]
- exponential hazard *λ*_*i,j*_(*t, τ*) = *pe*^*rτ*^, corresponding to a Gompertz distribution, often used to model of all-cause mortality.[52]

There are many other potential choices for *λ*_*i,j*_(*t, τ*). It may be useful to define the hazard rate piecewise over *t, τ*, or both, either piecewise constant or functions.[53] The choice of hazard function is highly dependent on the nature of the system being modelled and the available data sources.[54] Hazard rates can be estimated from processed survival data, typically after smoothing[55] or time-to-event distributions.[38, 56]

Hazard can be adjusted to reflect covariates; Cox proportional hazard ratios can be used to calculate *λ*_*i,j*_(*t, τ* | **x**) = *λ*_*i,j*_(*t, τ* |0)*e*^*β·***x**^ for some covariates **x** *∈* ℝ*n* and a baseline hazard function *λ*_*i,j*_(*t, τ* | 0).[44, 48] Constant hazard multipliers can be used as calibration targets, to maintain the shape of the distribution,[53] and hazards can be adjusted to reflect lead-time biases in the data (see Appendix C.1). With minor alterations, one can also introduce hazards which are dependent on the current state of the system; for example, this allows for modelling of infectious disease models. See Appendix C.3 for further details.

### 2.5 Example model

An example sojourn time density model is shown in Figure 2, demonstrating the impact of *t*- and *τ* -dependent hazard rates. This model has four states, *A, B, C, D*. The transitions out of A are *λ*_*A,B*_(*t, τ*) = *λ*_*A,C*_(*t, τ*) = 0.2, as these are constant, *P* (*X*_*t*_ = *A*) decays exponentially, like a Markov process (Figure 3). The subsequent transitions are *λ*_*B,D*_(*t, τ*) = 0 if *τ <* 1, 5 otherwise, and *λ*_*C,D*_(*t, τ*) = 0 if *t <* 2, 2 otherwise. The impact of the *t* and *τ* dependencies can be seen in the evolving density of *f*_*i*_(*t, τ*). The initial condition was *f*_*A*_(0, *τ*) = 1 for *τ ∈* [0, 1). Further examples are included in Appendix C.2.

**Figure 2.**
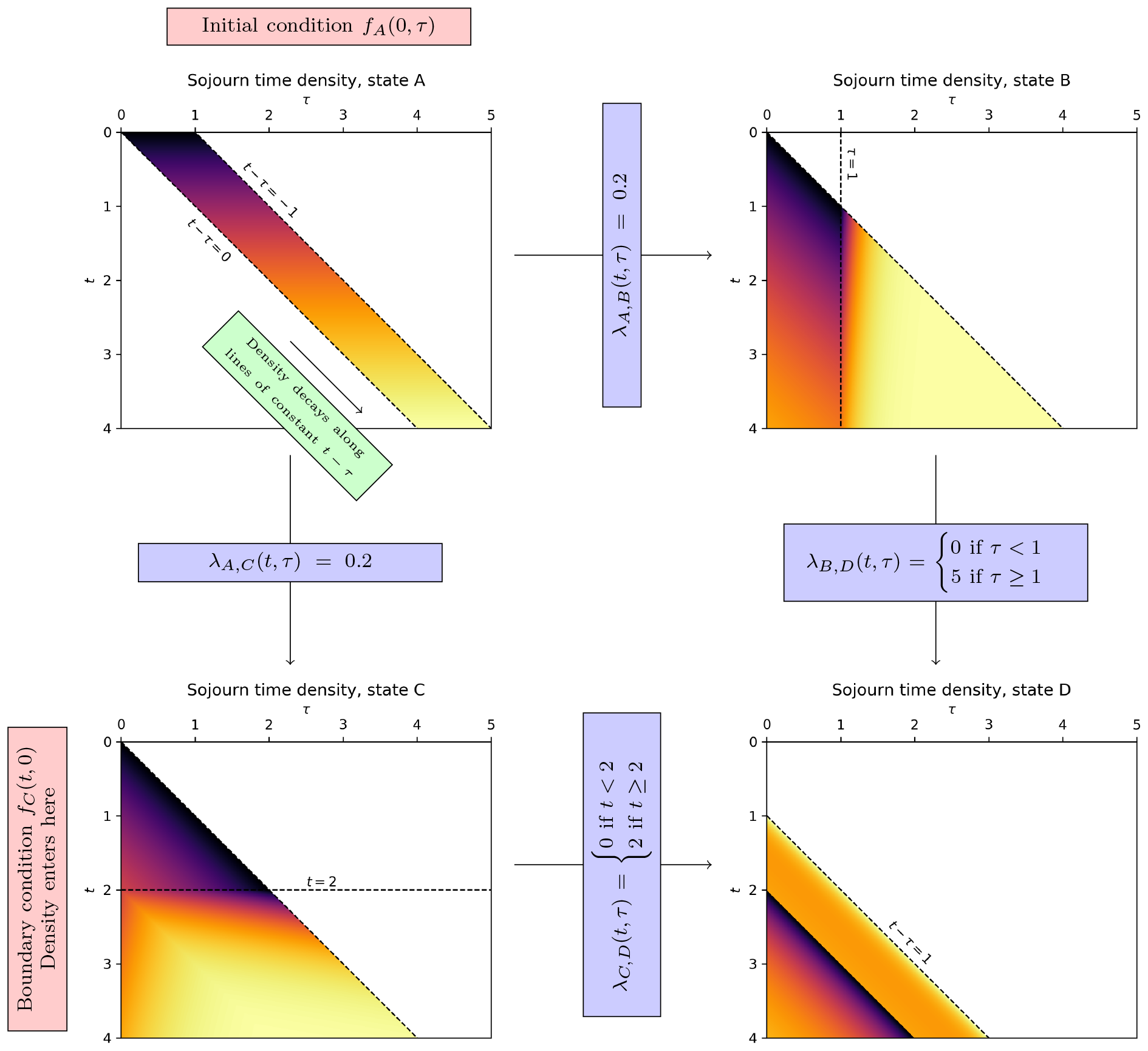
An example sojourn time density model, showing the evolution of the density of sojourn time. Darker regions have higher density, and lighter regions have lower density. Note how the density in state B sharply decreases at τ ≥ 1, and in state C at t ≥ 2. See Figure 3 for the resulting process.

**Figure 3.**
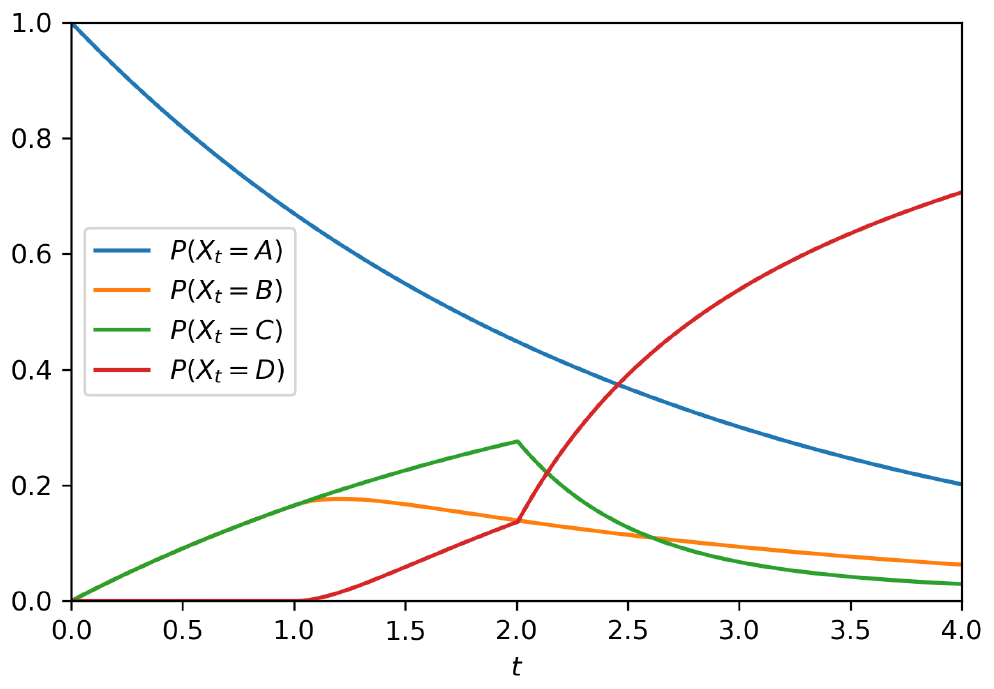
The resulting process X_t_ from the model illustrated in Figure 2.

## 3 Methods: Modelling Liver cirrhosis and HCC

We now describe the development of a sojourn time density model of liver disease and HCC. The Consolidated Health Economic Evaluation Reporting Standards (CHEERS)[57] checklist is included in Appendix D.

### 3.1 Model structure

The structure of the model is illustrated in Figure 4. Individuals start with compensated liver cirrhosis, which can develop into decompensated cirrhosis and/or undiagnosed HCC. These states were selected to capture the most clinically-relevant details based on the available data. Undiagnosed HCC is diagnosed either symptomatically/incidentally or at a routine HCC surveillance event. The Australian liver cirrhosis patient population was modelled.

**Figure 4.**
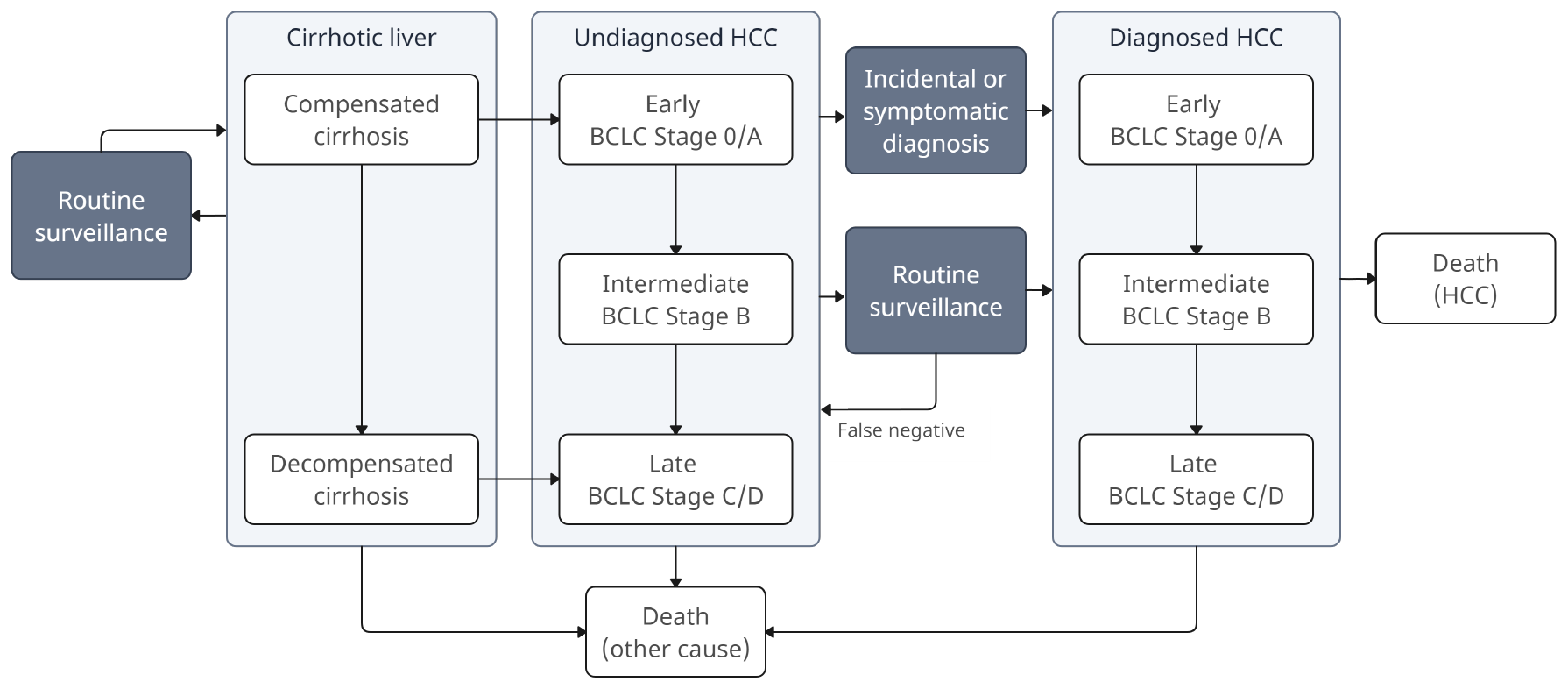
A simplified schematic of the model of HCC and liver surveillance.

HCC stage at diagnosis was modelled according to the Barcelona Clinic Liver Cancer (BCLC) staging system,[58] which is based on a patient’s Eastern Cooperative Oncology Group (ECOG) performance status,[59] Child-Pugh Score,[60] and tumour stage. Cancers were identified as early (BCLC stage 0/A: ECOG status 0, tumour *≤*3cm), intermediate (BCLC stage B: ECOG status 0, multinodular), or late stage (BCLC stage C/D: ECOG status *≥* 1, vascular invasion, extrahepatic spread, and/or Child-Pugh C). These groupings aligned with available survival data. Patients with compensated cirrhosis initially develop early stage HCC, which can subsequently progress; patients with decompensated cirrhosis develop late stage HCC only. The modelled stage is stage at diagnosis only; post-diagnosis progression is not explicitly modelled, but is reflected in modelling of stage-at-diagnosis survival per the available data.

### 3.2 Cirrhosis and health state transitions

Decompensation rates for patients with compensated liver cirrhosis were directly available from cohort studies.[61] HCC incidence, diagnosis, and upstaging rates were calibrated based on time-to-event data for HCC diagnosis and observed stage at diagnosis.[61, 62] This was supplemented with data on observed stage at diagnosis in cohorts routinely undergoing HCC surveillance[62] and data on the sensitivity and specificity of HCC surveillance.[63] The full list of sources for the model parameters are included in A.

### 3.3 HCC and other cause survival

Five-year stage-specific overall survival rates were calculated based on data from the NSW Cancer Registry, as these data are stratified by stage,[64] augmented by national data.[65] International studies were used to inform survival by time since diagnosis, by stage, and screen-vs symptomatically-detected HCC.[24] Treatment modalities, including serial treatments, were not modelled explicitly.

HCC recurrence over five years after initial diagnosis was not modelled due to lack of data; HCC recurrence within five years of the initial diagnosis is included in the modelled five-year survival rates. Other-cause mortality was modelled for people with compensated or decompensated cirrhosis [29] relative to the age-specific mortality rate in Australian data.[66]

### 3.4 Routine HCC surveillance

HCC surveillance is modelled as routine US with or without AFP after the diagnosis of compensated cirrhosis. Six-monthly US with AFP was modelled as the base case, as currently recommended.[8, 67, 68] Diagnosis to confirm suspected HCC, either after symptomatic development or positive surveillance event (including false positives), was modelled as computed tomography or magnetic resonance imaging, with biopsy in cases where imaging was insufficient.[10] People who experience liver decompensation do not recieve HCC surveillance, per recommendations.[7] For the purposes of this study, patients were assumed to have perfect uptake and adherence to surveillance, to assess the impact of the surveillance recommendations rather than of patient behaviours.

### 3.5 Costs and health state utilities

The benefits of surveillance were measured in quality-adjusted life-years. Disutilities were identified for patients with compensated cirrhosis, decompensated cirrhosis, and HCC patients. Disutilities for HCC patients were classified according to their phase of care (diagnostic, ongoing, or terminal), as well as multiplicative existing disutilities due to cirrhosis.[69, 70]

Costs and benefits were analysed from a health system perspective to capture the most relevant cost for Australian governmental policymakers, and presented as 2023 Australian dollars with costs adjusted using the Australian health CPI index.[71] Costs relating to cancer treatment were based on an excess-cost study which stratified patients by their primary treatment,[23] which varied by stage at diagnosis. These data were chosen as they are locally relevant and based on real-world observations rather than idealised treatment recommendations.

Costs for surveillance and diagnostic procedures were collated from the Australian Medicare Benefits Schedule. Other costs included annual costs of cirrhosis care for patients with and without decompensation, and end-of-life costs.[72, 73] A lifetime time horizon was included to capture the full downstream benefits of surveillance. For the cost-effectiveness analysis, a 5% annual discount rate was applied to both costs and QALYs, in line with other health economic analyses in Australia.[74] A supplementary sensitivity analysis on the impact of the choice of discount rate was completed. Cost inputs are included in Appendix A.

### 3.6 Parameters and calibration

Parameters were directly derived or calibrated from relevant sources identified in the clinical guidelines,[8] with Australian studies used where possible. Where Australian trial data were not available, the data sources prioritised meta-analyses with large cohort sizes.

Parametric forms for the hazard rates were chosen to best fit the data while minimising the number of parameters to avoid potential overfitting. Where necessary, the Nelder-Mead algorithm was used to determine the best-fit parameters by minimising the mean square error between the model outputs and the target data. Additional methodology is included in Appendix A.

### 3.7 Scenarios modelled

For the baseline analysis, we estimated the health benefits and economic impacts of 6-monthly HCC surveillance with US, with or without AFP, with no routine HCC surveillance as the comparator. This was modelled for a cohort with mean age 51 (standard deviation 11) to reflect the general cirrhotic population,[75] by aggregating single-age simulations. We calculated HCC stage, likelihood of HCC death, quality-adjusted life expectancy, and costs. A further analysis was conducted to assess the impact the frequency of surveillance.

### 3.8 Sensitivity analyses

One-way sensitivity analyses and probabilistic sensitivity analyses were conducted to assess the model’s sensitivity to key parameters. Additional methodology is included in Appendix B.

## 4 Results

### 4.1 Model calibration

The model was calibrated to reproduce key targets (Figure 5). The model was well-fitted to the target data, reproducing the terminal values for the survival curves and remaining within the 95% confidence intervals for the duration of the survival data. The full parameter set for the model is shown in Appendix A, including calibration targets and parameter distributions used in the sensitivity analyses.

**Figure 5.**
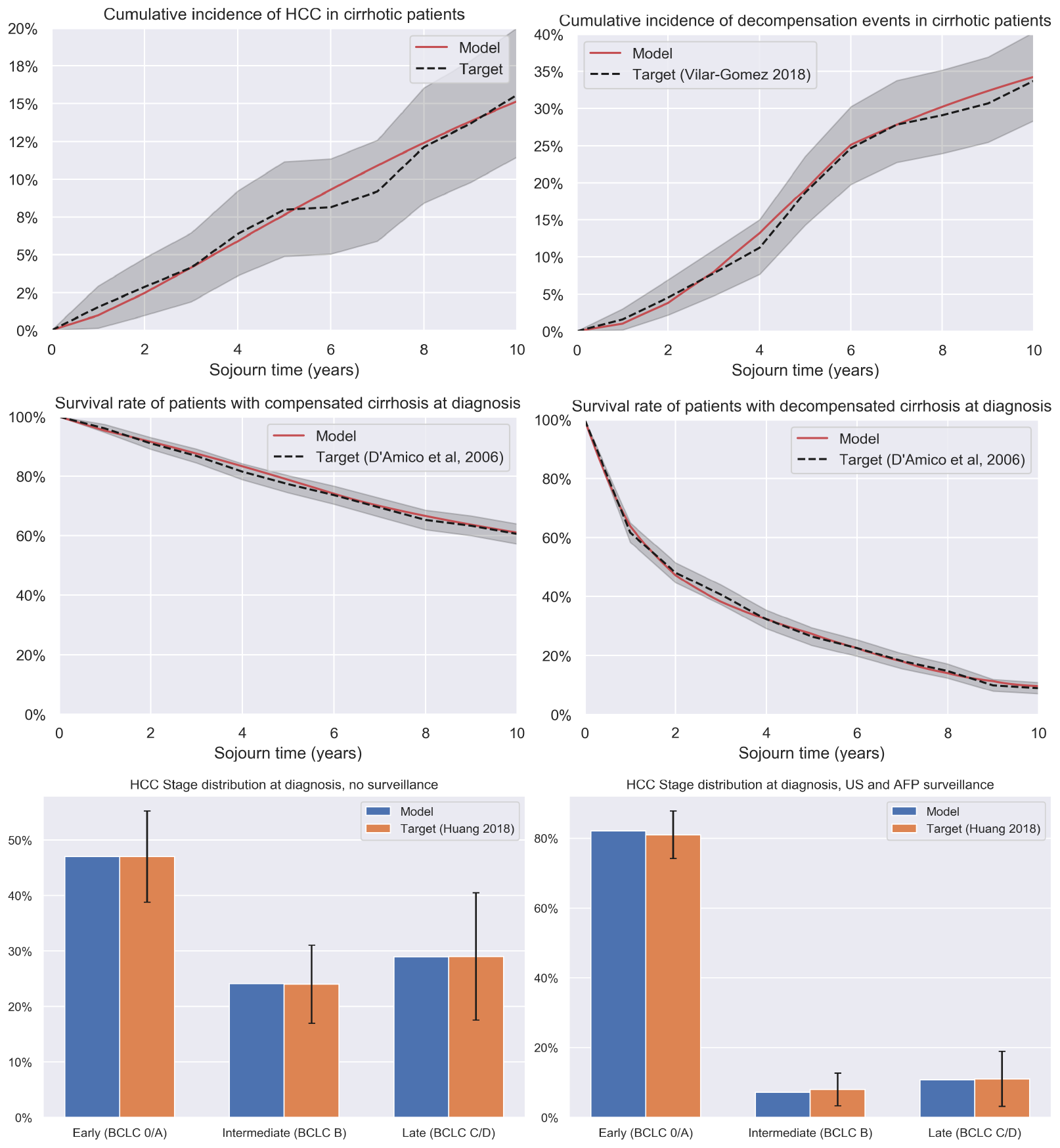
Key calibration outputs. From top to bottom: HCC incidence rates (left; target Vilar Gomez 2018 [61]) and decompensation rates (right); all-cause death rates in patients with compensated (left; includes any decompensation events) and decompensated (right) cirrhosis; HCC stage at diagnosis with (left) and without (right) routine HCC surveillance.

### 4.2 Health benefits and cost-effectiveness of surveillance

The results of the analysis are shown in Table 1. These analyses were for a cirrhotic cohort with no decompensation, and a mean age of 51 *±*11 years. Among those diagnosed with HCC, the probability of being diagnosed at early-stage disease would increase by 81.3% for those that undergo six-monthly US surveillance with AFP.

**Table 1:** Key results for the health benefits and cost-effectiveness of routine HCC surveillance by 6-monthly ultrasound vs no surveillance, with or without AFP. Costs include costs associated with ongoing cirrhosis care, surveillance costs, HCC diagnosis and treatment costs, and end-of-life costs. US - ultrasound. AFP - alpha-fetoprotein. HCC - hepatocellular carcinoma. CER - cost-effectiveness ratio vs no surveillance. ICER - incremental cost-effectiveness ratio vs previously most cost-effective strategy.

| Outcome | No Intervention | 6-monthly US | 6-monthly US with AFP |
| --- | --- | --- | --- |
| HCC incidence per 100,000 | 21,541 | - | - |
| Proportion of HCC diagnosed at early stage* | 47.0% | 79.6% | 81.3% |
| HCC mortality per 100,000 | 14,918 | 11,680 | 11,504 |
| Reduction vs no surveillance | - | 21.7% | 22.9% |
| Lifetime liver-related healthcare costs per 100,000 (billions) | \$12.91 | \$13.63 | \$13.68 |
| Increase in costs vs no surveillance | - | 5.6% | 5.9% |
| Liver costs per 100,000 (billions, discounted) | \$8.30 | \$8.89 | \$8.99 |
| Quality-adjusted life expectancy per 100,000 | 865,630 | 919,300 | 922,300 |
| Increase vs no surveillance | - | 6.2% | 6.5% |
| Quality-adjusted life expectancy per 100,000 (discounted) | 562,200 | 579,800 | 581,000 |
| CER vs No Intervention | - | \$33,850 | \$36,870 |
| ICER | - | \$33,850 | \$81,360 |

Six-monthly HCC surveillance with US alone would reduce a person with cirrhosis’ likelihood of HCC death by 21.7% compared to no intervention, increasing the quality-adjusted life-expectancy by 6.2%. Six-monthly US with the addition of AFP would reduce likelihood of HCC death by 22.9%, increasing the quality-adjusted life-expectancy by 6.5%. On average, people with cirrhosis would experience 17.6 surveillance events over their lifetime, and have an average total cirrhosis and HCC costs of 136,324 (2023 AUD), a 5.6% increase compared to those who do not undergo surveillance, with similar increases with the addition of AFP.

The cost-effectiveness of HCC surveillance with six-monthly US would be 33,850/QALY compared to no surveillance, below the indicative willingness-to-pay threshold of 50,000/QALY often used in Australia. This indicates that surveillance using six-monthly US would be cost-effective. Six-monthly surveillance with US and AFP would have a very similar cost-effectiveness of 36,860/QALY compared to no surveillance and be cost-effective. The incremental cost-effectiveness ratio of US with AFP vs US alone would be $81,360/QALY, indicating it is unlikely to be an incrementally cost-effective compared to offering US only.

The impact of varying the interval for routine US is shown in Figure 6. Shorter surveillance intervals have higher benefits, but at correspondingly higher costs. Surveillance intervals of 7 months or longer had cost-effectiveness ratio under 30,000/QALY for US with or without AFP. Additional benefits and costs are in very similar proportions at intervals over 12 months, while shorter intervals led diminishing benefits per cost increase. The addition of AFP was approximately equivalent to extending the surveillance interval by 1-3 months.

**Figure 6.**
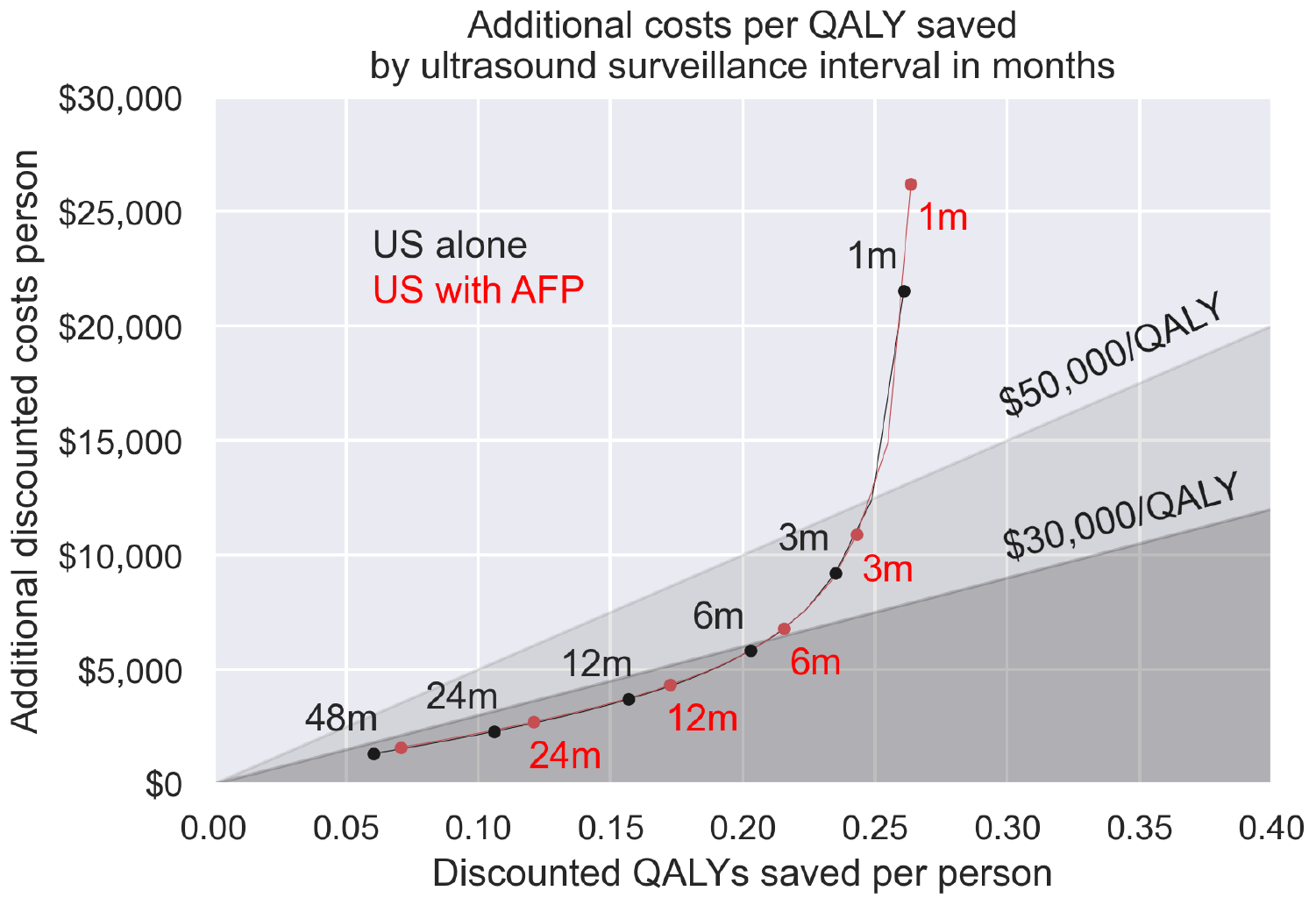
Discounted costs vs discounted QALYs saved for routine ultrasound surveillance with alpha-fetoprotein by surveillance interval in months, from 1 month to 48 months.

The results of the one-way sensitivity analysis found that the cost-effectiveness was most sensitive to parameters regarding early-stage HCC, including 5-year survival for Stage 0/A HCC and the proportion of HCCs diagnosed at Stage 0/A. The probabilistic sensitivity analysis found that six-monthly US was cost-effective in the 85.5% of simulations, demonstrating that this result is robust to parameter uncertainty. The analysis of the discount rate found that lower discount rates led to more favourable cost-effectiveness, due to the long-term benefits of surveillance. The full results are included in Appendix B.

## 5 Discussion

We described the development of a model of HCC surveillance in Australians with liver cirrhosis. This leverages a novel mathematical approach to simulating the sojourn time allowing detailed survival data sources to be used and short timescale events to be modelled. The model was calibrated to capture health outcomes and costs and reproduced the targets to a high level of precision.

Our findings indicate that routine HCC surveillance with six-monthly US with perfect adherence would be cost-effective and reduce the likelihood of HCC death. These results align with similar recent cost-effectiveness studies in Australia[10] and internationally[25, 8]. This analysis found that the addition of AFP would have further health benefits and would be cost-effective compared to no surveillance, but would not be incrementally cost-effective; there was a high degree of uncertainty around this outcome, as demonstrated by the included sensitivity analyses. Earlier versions of these findings were included in the *Clinical practice guidelines for HCC surveillance for people at high risk in Australia*[8] to support recommendations for the provision of US surveillance, which were endorsed by the Australian Government National Health and Medical Research Council in 2023.

We also analysed the impact of differing surveillance intervals, finding that routine HCC surveillance at an interval of 7 months or longer would be cost-effective under a $30,000/QALY threshold, while intervals of 3 months or longer would be cost-effective under a $50,000/QALY threshold. These findings indicate that the current six-month interval recommendation is appropriate, but also longer intervals would remain cost-effective and still have some health benefits. Longer surveillance intervals caused by patient noncompliance, delays, resource limitations, or other reasons would therefore remain cost-effective and have health benefits. There is limited longitudinal data on liver surveillance compliance with small sample sizes;[76] however, a correlation between compliance and HCC survival has been observed.[77] We also found that the addition of AFP may allow for similar benefits and costs as US alone at a longer interval.

The novel sojourn time density model structure allows for flexible analyses which rely on detailed survival data and flexible timescales. Most liver cancer models are Markov models,[8, 7] which may not be able to exploit complex survival data or reflect detailed disease progression.[78, 79] Liver disease states, both as clinically reported and as typically modelled, are usually a set of well-defined discrete disease states which serve as proxies for a more complex continuous spectrum of disease, such as HCC staging which simplifies variables such as the number and size of tumours, liver function, and other factors. Approaches to address this, like hidden states,[80] can increase the potential detail at the cost of interpretability. Modelling sojourn times based on survival data can reduce reliance on unobservable states and the reliance on detailed calibrations and potential for overfitting. By using the hazard rates as model parameters, the model can be interpreted directly in the context of source epidemiology studies, and parameter values can be directly inferred from survival data. Our approach to modelling also avoids relying on a fixed time discretisation, which could lead to numerical errors if chosen poorly[34] and can limit a model’s ability to capture short-timescale events such as late-stage HCC survival without recalibration. This may limit the model’s ability to reflect the true health benefits of early-stage diagnosis. Our model can be numerically evaluated at any timescale, allowing us to make simulate surveillance intervals which were not been included in real-world trials. Alternative model structures such as discrete event simulations[32, 33, 34], which are commonly used in cancer modelling, [18, 81, 82, 83, 84] agent-based/microsimulation models,[35, 14] and semi-Markov models[36, 37, 38] can allow for complex distributions of times between health states. However, these typically require stochastic evaluation,[38, 85, 86] which slows model performance and therefore limits the capacity for iterative model design and methods which explore wide parameter spaces, such as probabilistic sensitivity analysis. Designing models that reduce computational burden to facilitate probabilistic sensitivity analyses has been identified as a priority for health technology assessments,[87] with surrogate methods such as metamodelling reducing the reliance on larger models.[88, 89, 90] Our approach is fully deterministic and uses a simple numerical scheme, avoiding long runtimes as well as the need for sampling. It is also very flexible, with the capacity to capture a wide range of simulations, and can be expanded to capture systems such as infectious disease models as well (see Appendix C.3).

There is great potential to refine survival analysis estimates designed to guide policy recommendations,[91] and it is hoped that this modelling approach allows for the development of straightforward models which incorporate existing data sources in a clear and flexible way, reducing the need for model calibration and minimising data post-processing and model design effort. The benefits of this approach are demonstrated in the closeness-of-fit to the calibration targets (Figure 5) as well as the flexibility to evaluate surveillance at a wide range of intervals (Figure 6).

In future, this model will be used to analyse combinations of surveillance technologies and intervals. This will allow us to analyse more complex surveillance algorithms, such as the FIB-4 algorithm recommended by the Asian Pacific Association for the Study of the Liver clinical guidelines,[92] and optimise these algorithms through iterative design enabled by the fast computation time and high precision.

The limitations of this model structure include the semi-Markov property; each state transition depends only on the current state, the sojourn time, and the model time. This approach may therefore not be appropriate in contexts where multiple prior events are required to inform future likelihoods, with the number of states needed to represent these growing exponentially.

This model considers only the Australian population with liver cirrhosis. Liver cirrhosis is typically attributable to chronic hepatitis B, chronic hepatitis C, alcohol-related liver disease, metabolic-associated fatty liver disease, or a combination of these. Each of these groups have differing HCC risks, potentially differing treatment needs,[93] and differing prevalence in the Australian population.[11, 94] Although these groups are included in the aggregate liver cirrhosis population modelled here, they are not modelled explicitly by aetiology. Future model expansions to capture these risk groups explicitly, will allow more precise cost-effectiveness estimates and recommendations by group, identifying the optimal benefit of health benefits and costs between surveillance technology options and varying intervals.

Another limitation of our model is that treatment is not explicitly modelled by modality; patients are instead modelled based on aggregate survival and costs by stage at diagnosis. Although this is in line with many other models of cancer and surveillance,[15] models of HCC treatment can provide additional detail, particularly regarding health service utilisation.[10] Another limitation is the uncertainty around appropriate willingness-to-pay thresholds for Australia, with both 30,000 and 50,000 per QALY used. The interpretation of these results should consider the context to determine the apppriate threshold.

The current simulation of the influence of age on HCC risk was limited by a lack of data. Currently, the modelled age influences HCC risk through time exposed to cirrhosis and risk of other-cause mortality. Incorporating further targets for the evolving risk of HCC and stage at diagnosis would improve the cost-effectiveness estimates and help refine recommendations. In general, data regarding HCC development and surveillance is limited; for example, differences in estimated stage at diagnosis,[62, 95] with significant differences likely attributable to surveillance adherence and patient cohorts, and sensitivity and specificity of US.[96, 63] As further data are made available, modelling will be updated and refined to help guide interventions.

## 6 Conclusions

There is great potential for routine HCC surveillance to improve mortality outcomes in Australia. However, any recommendations must account for the costs and the consequences of the lower life expectancy and quality-of-life for high-risk individuals. Our robust and flexible model of liver disease and surveillance can estimate the impact of surveillance interventions and inform planning and policy in this area. Future evaluations will provide refinements and extensions of the economic evaluations to capture detailed patient risk groups and alternative surveillance modalities.

## Data Availability

All data produced in the present work are contained in the manuscript. The modelling in this manuscript is informed by the parameters listed in the appendix.

## Acknowledgements

The authors would like to acknowledge the Expert Advisory Group for the Optimising Liver Cancer Control in Australia convened by Cancer Council Australia. We would also like to thank Cancer Council Australia for coordination of this project.

Materials reported in this manuscript, including the modelling and results, were developed to support the Optimising Liver Cancer Control in Australia project and were originally developed and reported in *[Clinical practice guidelines for hepatocellular carcinoma surveillance for people at high risk in Australia: Appendix D7](https://www.cancer.org.au/clinical-guidelines/liver-cancer/hepatocellular-carcinoma)*, a non-peer reviewed report.

The authors would like to acknowledge the Working Group and Community Reference Group for the Clinical practice guidelines for hepatocellular carcinoma surveillance for people at high risk in Australia. A full list of Working Group members is available in *[Clinical practice guidelines for hepatocellular carcinoma surveillance for people at high risk in Australia: Appendix I](https://www.cancer.org.au/clinical-guidelines/liver-cancer/hepatocellular-carcinoma)*.

The authors would like to thank the Daffodil Centre Systematic Review team, who provided assistance in reviewing the literature on existing models of HCC and surveillance, as well as Yue He, who gave valuable feedback on the manuscript.

This manuscript uses custom data sets from the Australian Institute of Health and Welfare and the Cancer Institute NSW. We thank the Australian Institute of Health and Welfare and the population-based cancer registries of New South Wales, Victoria, Queensland, Western Australia, South Australia, Tasmania, the Australian Capital Territory and the Northern Territory for the provision of data from the Australian Cancer Database. We would also like to thank the Cancer Institute NSW for their assistance with the data request.

Karen Canfell receives salary funding from the National Health and Medical Research Council Australia (NHMRC Leadership Fellowship APP1194679).

## Footnotes

1 We refer to *survival* in the sense of survival analysis, i.e. the time until an event of interest,[39] not necessarily mortality

