## Appendices A-D for "An economic evaluation of routine hepatocellular carcinoma surveillance for high-risk patients using a novel approach to modelling competing risks"

### Appendix A Model calibration and parameters

For each transition, appropriate target data were identified. Data sources most likely to reflect the progression of liver disease in the Australian population were prioritised wherever possible. The model was then calibrated to reproduce these targets. For data depending on sojourn time, including time to liver decompensation, time to death in patients with cirrhosis, and survival time in those diagnosed with HCC, the hazard was fitted to a function chosen to match the data.

The parameters and calibration data are shown in Tables A1,A2,A3,A4,A5,A6. Also included are parameter distributions used in the one-way and probabilistic sensitivity analyses (see B). The parameter distributions were calibrated to reproduce the statistical confidence intervals reported in the original data sources wherever possible, with exceptions noted. Relative hazards were modelled as lognormally distributed, to align with the log-log survival estimates used to inform the parameters.[1] Proportions, including disutilities, were modelled as beta distributions.[2] Costs were also modelled as lognormally distributed.

*Table A1: Calibrated natural history parameters for HCC.*

| **Parameter** | **Model value** | **Target** | **Source/notes** |
|---|---|---|---|
| Cirrhosis decompensation hazard | $\lambda = \begin{cases} 0.0204\tau + 0.0106 \text{ if } \tau < 6 \\ 0.0554 \text{ if } \tau \geq 6 \end{cases}$ [1] | | Calibrated to reproduce rates in [3] (Figure 5); examples below.[2] |
| Annual decompensation risk | 1.03% | 1.59% (0.17-3.01%) | Calibration target [3]. |
| Ten-year decompensation risk | 34.5% | 33.7% (28.4-39.1%) | |
| HCC onset hazard | $\lambda = 0.0102\tau^{1.377}$ | | Calibrated to reproduce rates in [3] (Figure 5); examples below.[2] |
| Annual HCC risk | 0.99% | 1.53% (0.14-2.93%) | Calibration target [3]. |
| Ten-year HCC risk | 15.2% | 15.6% (11.5-19.7%) | |
| Death hazard (compensated) | $\lambda = 0.0336\tau^{-0.2379}$ | | Calibrated to reproduce rates in [4] (Figure 5); examples below.[2] |
| Annual death risk (compensated) | 4.72% | 3.97% (2.62-5.31%) | Calibration target [4]. |
| Ten-year death risk (compensated) | 39.4% | 39.4% (36.1-42.8%) | |
| Death hazard (decompensated) | $\lambda = (0.1539\tau - 0.4865)^2 + 0.2103$ | | Calibrated to reproduce rates in [4] (Figure 5); examples below.[2] |
| Annual death risk (decompensated) | 37.8% | 38.3% (35.0-41.6%) | Calibration target [4]. |
| Ten-year death risk (decompensated) | 90.6% | 91.1% (89.2-93.0%) | |
| HCC progression hazard, BCLC A | 0.2049 | | Calibration target for stage at diagnosis (see Table A3). |
| HCC symptomatic/incidental detection hazard, BCLC A | 0.2091 | | |
| HCC progression hazard, BCLC B | 1.0259 | | As above |
| HCC symptomatic/incidental detection hazard, BCLC B | 1.1727 | | |
| HCC symptomatic/incidental detection hazard, BCLC C/D | 1.4887 | | |
| HCC five-year survival, BCLC 0/A/B | 47.7% | 47.7% | Calibration target, NSW Cancer Registry[3,4] |
| HCC five-year survival, BCLC C/D | 20.6% | 20.6% | As above |
| HR for survival, stage B vs 0/A | 0.508 | 0.508 | Calibration target [5]. |
| HR for survival, stage D vs C | 0.841 | 0.841 | |

[1] $\tau$ is the sojourn time
[2] The full calibration included a range of target data points not shown here.
[3] NSW Cancer Registry. Data request: Hepatocellular carcinoma (coded as C22.0 or C22.9) incidence and mortality). Private communication; unpublished.; 2021 Aug.
[4] Additional data was used to calibrate detailed survival by time since diagnosis.[5]

*Table A2: Parameter distributions for probabilistic sensitivity analysis: liver disease and HCC parameters.*

| **Parameter** | **PSA Distribution** | **Source/notes** |
|---|---|---|
| Relative risk of cirrhosis decompensation[1] | Lognormal($\mu = -.007, \sigma = .117$) | [3] |
| Relative risk of HCC development[1] | Lognormal($\mu = -.013, \sigma = .158$) | |
| Relative risk of death (compensated cirrhosis) | Lognormal($\mu = -.011, \sigma = .148$) | [4] |
| Relative risk of death (decompensated cirrhosis) | Lognormal($\mu = -.004, \sigma = .09$) | |
| Relative risk of stage progression, undetected stage 0/A HCC | Lognormal($\mu = -.017, \sigma = .185$) | Calibration target; see Table A3. |
| Relative risk of detection, undetected stage 0/A HCC | Lognormal($\mu = -.017, \sigma = .183$) | |
| Relative risk of stage progression, undetected stage B HCC | Lognormal($\mu = -.057, \sigma = .336$) | |
| Relative risk of detection, undetected stage B HCC | Lognormal($\mu = -.054, \sigma = .328$) | |
| Relative risk of detection, undetected stage C/D HCC | Lognormal($\mu = -.001, \sigma = .051$) | |
| Relative risk HCC death, stage 0/A HCC | Lognormal($\mu = -.011, \sigma = .149$) | [5] |
| Relative risk HCC death, stage B HCC | Lognormal($\mu = -.006, \sigma = .108$) | |
| Relative risk HCC death, stage C/D HCC | Lognormal($\mu = -.003, \sigma = .081$) | |

[1] Relative risk vs baseline rates in Table A1.

*Table A3: Calibrated HCC parameters and targets.*

| Parameter | Model value | Target | PSA distribution | Source/notes |
|---|---|---|---|---|
| Ultrasound sensitivity (stage 0/A) | 53% | 53% (35-70%)[1] | Beta($\alpha = 15.87, \beta = 14.23$) | Calibration target [6]. |
| Ultrasound sensitivity (stage B/C/D) | 84% | 84% (67-92%) | Beta($\alpha = 29.03, \beta = 6.74$) | |
| Ultrasound specificity | 91% | 91% (86-94%) | Beta($\alpha = 186.8, \beta = 19.86$) | |
| Ultrasound & AFP sensitivity (stage 0/A) | 63% | 63% (48-75%) | Beta($\alpha = 29.69, \beta = 18.22$) | Calibration target [6]. |
| Ultrasound & AFP sensitivity (stage B/C/D) | 97% | 97% (91-99%) | Beta($\alpha = 84.54, \beta = 3.53$) | |
| Ultrasound & AFP specificity | 84% | 84% (77-89%) | Beta($\alpha = 121.2, \beta = 24.07$) | |
| Stage 0/A HCC, no surveillance[3] | 47% | 47% | | Calibration target [7]. |
| Stage B HCC, no surveillance | 24% | 24% | | |
| Stage C/D HCC, no surveillance | 29% | 29% | | |
| Stage 0/A HCC, US surveillance[4] | 82% | 81% | | |
| Stage B HCC, US surveillance | 7% | 8% | | |
| Stage C/D HCC, US surveillance | 11% | 11% | | |
| Stage 0/A HCC, US & AFP surveillance[3] | 83% | - | | Model outcome.[3] |
| Stage B HCC, US & AFP surveillance | 7% | - | | |
| Stage C/D HCC, US & AFP surveillance | 10% | - | | |

[1] Reported 95% confidence interval.
[2] With six-monthly surveillance.
[3] BCLC stage at diagnosis. Excluding any post-diagnosis stage progression.
[4] Based on HCC progression and surveillance test characteristics.

*Table A4: Treatment modality proportions for diagnosis and treatment of HCC.*

| **Parameter** | **Model value** | **PSA Distribution** | **Source/notes** |
|---|---|---|---|
| HCC diagnosis[1]: CT[2] | 80% | Beta($\alpha = 163.98, \beta = 23.94$) | Source: [8]. |
| HCC diagnosis: MRI[3] | 20% | Beta($\alpha = 23.12, \beta = 161.89$) | |
| HCC diagnosis: Biopsy[4] | 10% | Beta($\alpha = 18.13, \beta = 165.82$) | |
| Stage 0/A treatment[5]: transplant | 19.0% | Beta($\alpha = 12.86, \beta = 54.81$) | Source: [9]. |
| Stage 0/A treatment: resection | 13.8% | Beta($\alpha = 3.05, \beta = 19.04$) | |
| Stage 0/A treatment: ablation | 25.6% | Beta($\alpha = 16.48, \beta = 47.91$) | |
| Stage 0/A treatment: TACE | 34.8% | Beta($\alpha = 21.33, \beta = 39.96$) | |
| Stage 0/A treatment: resection, ablation/TACE, sorafenib | 3.4% | Beta($\alpha = 3.05, \beta = 86.61$) | |
| Stage 0/A treatment: ablation, sorafenib | 1.5% | Beta($\alpha = 1.45, \beta = 95.51$) | |
| Stage 0/A treatment: TACE, sorafenib | 2.0% | Beta($\alpha = 1.9, \beta = 93.19$) | |
| Stage B treatment: transplant | 8.3% | Beta($\alpha = 1.57, \beta = 17.37$) | |
| Stage B treatment: resection | 8.3% | Beta($\alpha = 1.57, \beta = 17.37$) | |
| Stage B treatment: ablation | 17.7% | Beta($\alpha = 2.77, \beta = 12.9$) | |
| Stage B treatment: TACE | 24.0% | Beta($\alpha = 3.44, \beta = 10.89$) | |
| Stage B treatment: ablation, sorafenib | 14.1% | Beta($\alpha = 2.38, \beta = 14.52$) | |
| Stage B treatment: TACE, sorafenib | 19.2% | Beta($\alpha = 2.95, \beta = 12.4$) | |
| Stage B treatment: resection, sorafenib | 8.3% | Beta($\alpha = 1.57, \beta = 17.37$) | |
| Stage C/D treatment: ablation | 3.4% | Beta($\alpha = 1.44, \beta = 41.00$) | |
| Stage C/D treatment: TACE | 4.6% | Beta($\alpha = 1.87, \beta = 38.68$) | |
| Stage C/D treatment: ablation, sorafenib | 6.8% | Beta($\alpha = 2.55, \beta = 34.93$) | |
| Stage C/D treatment: TACE, sorafenib | 9.2% | Beta($\alpha = 3.24, \beta = 31.99$) | |
| Stage C/D treatment: ablation, palliation | 11.9% | Beta($\alpha = 4.02, \beta = 29.79$) | |
| Stage C/D treatment: TACE, palliation | 16.1% | Beta($\alpha = 5.14, \beta = 26.77$) | |
| Stage C/D treatment: sorafenib, palliation | 16.0% | Beta($\alpha = 5.00, \beta = 26.25$) | |
| Stage C/D treatment: palliation | 8.3% | Beta($\alpha = 8.68, \beta = 18.44$) | |

[1] Including after a false positive surveillance event.
[2] CT: computed tomography.
[3] MRI: magnetic resonance imaging.
[4] After inconclusive CT or MRI.
[5] Treatments listed sequentially, with primary treatment modality listed first.

*Table A5: Cost parameters.*

| Item | Cost | PSA Distribution | Source/notes |
|---|---|---|---|
| Annual compensated cirrhosis care | $4,713 | Lognormal($\mu = 8.047, \sigma = 0.527$) | [10] |
| Annual decompensated cirrhosis care | $22,701 | Lognormal($\mu = 9.861, \sigma = 0.306$) | |
| Ultrasound | $115.75 | Lognormal($\mu = 4.731, \sigma = 0.104$) | Medicare benefits schedule.[2] |
| Alpha-fetoprotein | $24.35 | Lognormal($\mu = 3.172, \sigma = 0.104$) | |
| GP visit | $39.75 | Lognormal($\mu = 3.662, \sigma = 0.104$) | |
| CT | $499.50 | Lognormal($\mu = 6.193, \sigma = 0.104$) | |
| MRI | $558.80 | Lognormal($\mu = 6.305, \sigma = 0.103$) | |
| Liver biopsy | $377.20 | Lognormal($\mu = 5.912, \sigma = 0.104$) | |
| Transplant[3] | $320,107 | Lognormal($\mu = 12.731, \sigma = 0.367$) | [11] |
| Resection | $73,310 | Lognormal($\mu = 11.09, \sigma = 0.556$) | |
| Ablation (RFA/MWA/PEI)[4] | $94,611 | Lognormal($\mu = 11.292, \sigma = 1.168$) | |
| TACE[5] | $76,482 | Lognormal($\mu = 11.252, \sigma = 0.85$) | |
| Sorafenib | $42,338 | Lognormal($\mu = 10.561, \sigma = 0.622$) | |
| Palliation | $45,084 | Lognormal($\mu = 5.912, \sigma = 0.104$) | |
| HCC death costs | $44,945 | Lognormal($\mu = 10.713, \sigma = 0.011$) | [12] |
| Other cause death costs | $31,513 | Lognormal($\mu = 10.358, \sigma = 0.012$) | |

[1] Calibrated to ±20% costs, in the absence of appropriate targets.
[2] Department of Health, Australian Government. MBS Online: Medicare Benefits Schedule. [Internet]. Canberra, ACT: Commonwealth of Australia.
[3] Patient treatment costs are overall costs classified according to their primary treatment, following the methodology in Hong et al.[11] Patients may have further treatments -these costs are included in the figures presented.
[4] Proportion of patients allocated to RFA/MWA/PEI based on the proportions reported in Hong et al.[11]
[5] Including TACE with cisplantin, TACE with doxorubicin, and SIRT.

*Table A6: Health utility loss parameters for patients with cirrhosis and/or HCC.*

| **Health state** | **Disutility** | **PSA Distribution** | **Source/notes** |
|---|---|---|---|
| Annual disutility:[1]compensated cirrhosis | 0.32 | Beta($\alpha = 2664.42, \beta = 5662.60$) | [13] |
| Annual disutility: decompensated cirrhosis | 0.38 | Beta($\alpha = 864.17, \beta = 1411.13$) | |
| HCC, diagnostic phase, compensated cirrhosis[2] | 0.52 | Beta($\alpha = 101.97, \beta = 93.86$) | [14, 13][3] |
| HCC, controlled phase, compensated cirrhosis[4] | 0.35 | Beta($\alpha = 1573.04, \beta = 2860.77$) | |
| HCC, terminal phase, compensated cirrhosis[5] | 0.69 | Beta($\alpha = 50.00, \beta = 22.81$) | |
| HCC, diagnostic phase, decompensated cirrhosis | 0.76 | Beta($\alpha = 114.90, \beta = 88.87$) | |
| HCC, controlled phase, decompensated cirrhosis | 0.41 | Beta($\alpha = 2243.60, \beta = 3201.66$) | |
| HCC, terminal phase, decompensated cirrhosis | 0.72 | Beta($\alpha = 59.50, \beta = 23.85$) | |

[1] Annual utility loss vs mean utility for general population.

[2] "Diagnostic phase" includes any time in the first year after diagnosis (excluding time in terminal phase).

[3] Including disutilities from both compensated cirrhosis and cancer treatment, with multiplicative disutilities as described in [14].

[4] "Controlled phase" includes any time after the first year after diagnosis (excluding time in terminal phase).

[5] "Terminal phase" includes any time in the final year before HCC death.

### Appendix B Sensitivity Analyses

#### B.1 One-way sensitivity analysis

A one way sensitivity analysis was completed on the natural history parameters, using the 6 monthly ultrasound scenario. The parameter ranges were based on the 95% confidence intervals of the distributions for each parameter (see Tables A2, A3, A4, A5, A6 ). We found that the model results were robust to the key health parameters, and were most sensitive to parameters around early-stage (BCLC A/0) HCC. However, in all analyses the CER for 6-monthly US HCC surveillance remained below the $50,000/QALY threshold. The parameters with the greatest impact on the CER are shown in Figure B.7.

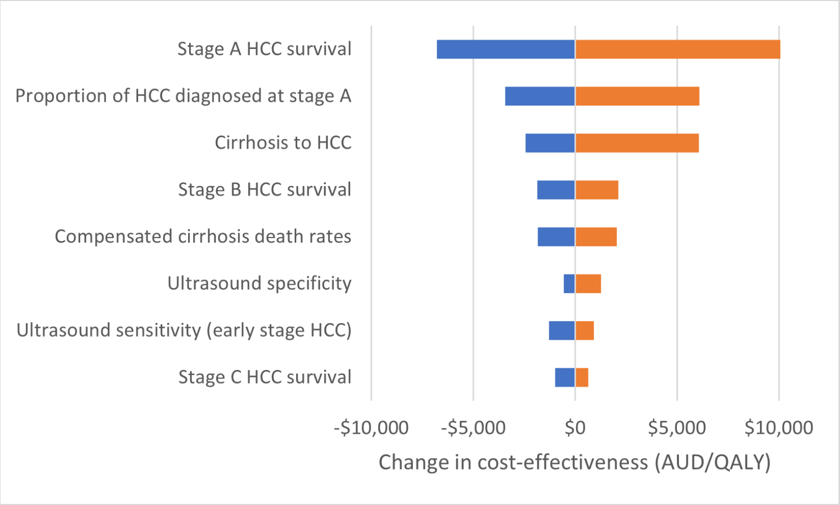


*Figure B.7: One-way sensitivity analysis results showing the impact of key model parameters on the cost-effectiveness of 6-monthly HCC surveillance with ultrasound.*

#### B.2 Discount rates

As the use of discounting in health economics varies by setting, it is important to understand the impact of this parameter choice. Although a 5% discount rate is typically used in Australia, there is some disagreement on this point. For example, Medicines Australia has suggested the use of a 1.5% discount rate,[15, 16] to emphasise the long-term impacts of an intervention.

To address this, a sensitivity analysis on the discount rates was completed on the discount rates used. We calculated the cost-effectiveness of US or US and AFP routine HCC surveillance with discount rates of 0%, 1.5%, 3%, 5%, and 7%, based on the most commonly used discount rates. The results of this analysis are shown in Table B7.

This analysis found that lower discount rates led to more favourable cost-effectiveness ratios. No discounting (0%) leading to a CER of $12,800 for US alone and $14,400 for US with AFP, increasing to $47,000 and $51,300 respectively for a 7% discount rate. Notably, the ICER for US with AFP vs US was $39,700 for a 0% discount rate, under a $50,000/QALY threshold. These findings reflect the fact that the cost of surveillance is a short-term additional cost, whereas the benefits are long-term life-expectancy improvements across a patient lifetime. Therefore any discounting reduces the percieved benefits and thus the CER.

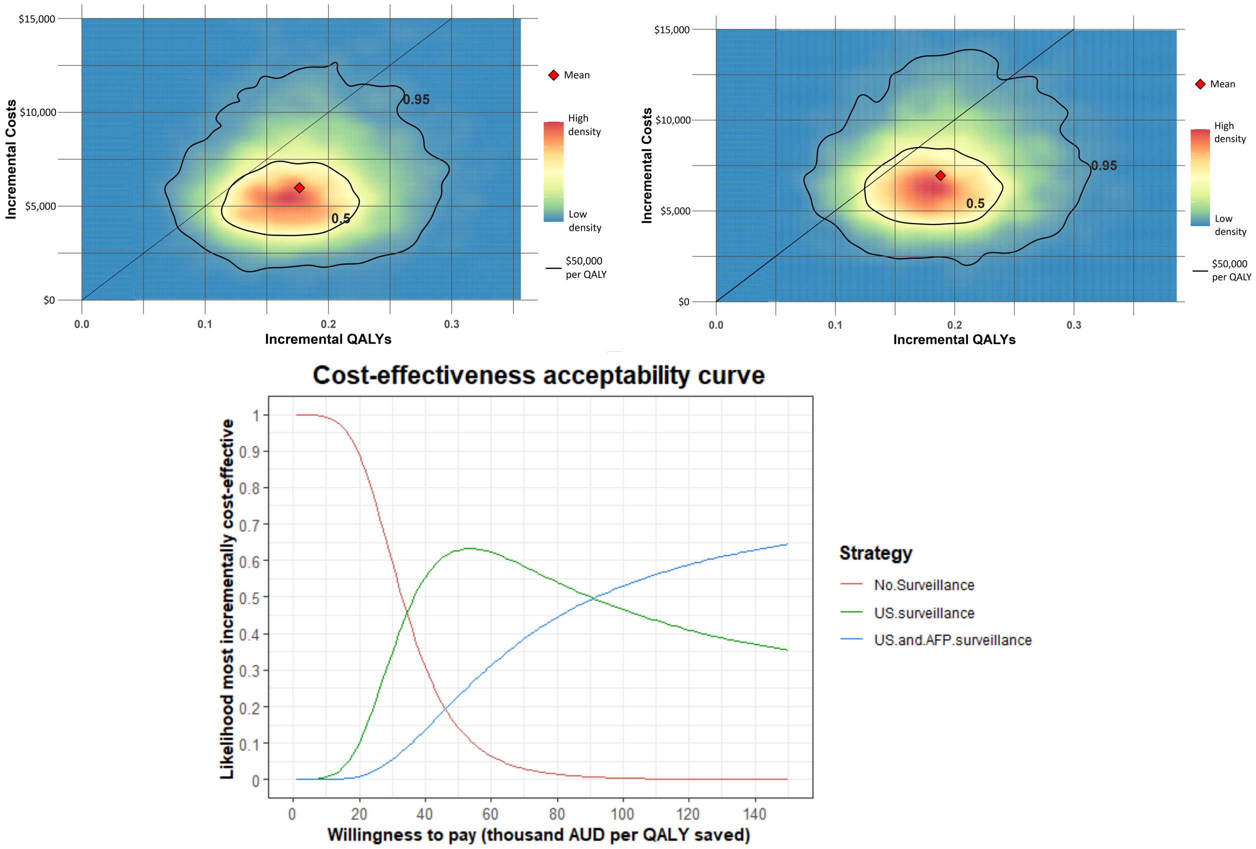


*Figure B.8: Results of the probabilistic sensitivity analysis.* **Top:** *relative density plot[17] of the additional costs and QALYs per person for 6-monthly HCC surveillance with ultrasound alone (left) and ultrasound and alpha-fetoprotein (right).* **Bottom:** *the incremental cost-effectiveness acceptability curve[18] showing the likelihood that no surveillance, ultrasound surveillance, or ultrasound and alpha-fetoprotein surveillance are the most incrementally cost-effective option at a range of willingness-to-pay thresholds, based on the proportion of simulations where each option is most cost-effective.*

*Table B7: Sensitivity analysis on the discount rate used. Discounting was applied to both QALYs and costs from surveillance start time.*

| | **No Surveillance** | | **US Surveillance** | | | **US + AFP Surveillance** | | | |
|---|---|---|---|---|---|---|---|---|---|
| **Discount rate** | Discounted costs per person | Discounted QALYs per person | Discounted costs per person | Discounted QALYs per person | CER vs no surveillance | Discounted costs per person | Discounted QALYs per person | CER vs no surveillance | ICER vs US alone |
| 0% | $129,000 | 8.65 | $136,000 | 9.15 | $12,800 | $137,000 | 9.18 | $14,400 | $39,700 |
| 1.5% | $111,000 | 7.46 | $118,000 | 7.82 | $17,500 | $119,000 | 7.84 | $19,500 | $50,800 |
| 3% | $97,400 | 6.54 | $104,000 | 6.81 | $23,400 | $105,000 | 6.82 | $25,900 | $64,000 |
| 5% | $83,000 | 5.62 | $88,900 | 5.8 | $33,900 | $89,900 | 5.81 | $36,900 | $81,400 |
| 7% | $72,000 | 4.93 | $77,600 | 5.05 | $47,000 | $78,600 | 5.06 | $51,300 | $116,000 |

US - ultrasound.
AFP - alpha-fetoprotein.
CER - cost-effectiveness ratio vs no surveillance.
ICER - incremental cost-effectiveness ratio vs previously most cost-effective strategy.

#### B.3 Probabilistic sensitivity analysis

A probabalistic sensitivity analysis (PSA) was completed to reflect the parameter uncertainty from the various sources. Parameter values were sampled from the distributions listed in Appendix A, and for each parameter set the model was run to simulate outcomes with no surveillance and 6-monthly HCC surveillance through US alone or US with AFP. The PSA simulated 50,000 parameter sets.

In 85.5% of simulated runs, 6-monthly HCC surveillance with US alone was cost-effective under the $50,000/QALY threshold, indicating the cost-effectiveness was robust to most parameter uncertainty. With the addition of AFP, 80.8% of simulated runs were cost-effective. The results of the PSA are illustrated in Figure B.8.

### Appendix C Additional notes and examples on sojourn density models

#### C.1 Initial conditions and lead time biases in sojourn time density models

The appropriate initial conditions for a sojourn time density model are highly dependent on the system being modelled. The Dirac delta function around $\tau = 0$ is a potential initial condition, corresponding to starting in state $i$ with no existing sojourn time; explicitly,

$$f_j(0,\tau) = \begin{cases} \delta(\tau) & \text{if } i = j \\ 0 & \text{if } i \neq j. \end{cases} \tag{33}$$

In practice, modelling may start 'in-situ', i.e. after some sojourn time, informing a distribution for the initial condition. For example, this could reflect the time an individual would have spent in a health state at the start of a trial. This may be equivalent to the lead time bias (Section C.1).

As the sojourn time is modelled explicitly in the sojourn time density framework, this allows us to incorporate lead time bias. Lead time biases occur in many contexts in epidemiology; for instance, early detection of a cancer artificially increases perceived survival times due to the increased observation window.[19, 20]

A related bias occurs in modelling which has been informed by survival data from trial outcomes. Real world survival data is measured from the start of a trial after diagnosis, which in turn occurs after asymptomatic onset of disease. However, epidemiological models typically use data from trial start time, in the absence of data from disease onset.

These biases can be addressed by modifying the hazard rate. A simple approach may be to define

$$\lambda_{i,j}(t,\tau) = \begin{cases} 0 & \text{if } \tau < \tau_0 \\ \tilde{\lambda}_{i,j}(t,\tau - \tau_0) & \tau \geq \tau_0 \end{cases} \tag{34}$$

where $\tilde{\lambda}_{i,j}(t,\tau)$ is the observed cause-specific hazard rate from a trial and $\tau_0$ is some cut-off time. In practice, an estimate for $\tau_0$ may be difficult to determine, requiring a sensitivity analysis over a range. A linear/sigmoidal interpolation between $\tau = 0$ and $\tau = \tau_0$ may also be useful, or a more nuanced approach using a Bayesian prior for the lead-time.[21, 22, 23]

We can also introduce transitions which in some sense "preserve" the sojourn time. For example, in an epidemiological model this may represent a transition which is independent of the underlying growth of disease, such as entry into a routine surveillance program; see Appendix C.3.

#### C.2 Example models

Further example sojourn time density models are shown in Figure C.9, to demonstrate potential approaches to designing sojourn time density models. In all examples the initial condition is a Dirac delta function in state $A$ at $\tau = 0$.

- **Figure C.9, top:** constant hazards
  - Four states $A$,$B$,$C$,$D$ with transitions $A \to B$, $B \to C$, $C \to D$.
  - Constant hazard rates $\lambda_{A,B}(t,\tau) = 1, \lambda_{B,C}(t,\tau) = 2, \lambda_{C,D}(t,\tau) = 3$.
  - As all hazard rates are constant, the resulting process $X_t$ is *Markov*.
  - For an appropriate discretisation $\Delta t$, this is equivalent to a standard *Markov model*.
- **Figure C.9, middle:** Model loops leading to oscillatory behaviour.

- Two states $A$,$B$ with transitions $A \to B$, $B \to A$.
- Weibull hazard rates for $\lambda_{A,B}(t,\tau) = 10\tau^9, \lambda_{B,A}(t,\tau) = 10\tau^9$.
- This is an example of a model with no terminal state; instead, the density oscillates from $A$ to $B$ and back, before reaching an equilibrium at $P(X_t = A) = P(X_t = B) = 0.5$.
- Simple models can lead to interesting dynamics.

- **Figure C.9, bottom:** Piecewise constant.
  - Two states $A$,$B$ with only one transitions $A \to B$.
  - The dots shown come from a (fictitious) Kaplan-Meier analysis.
  - The hazard rate $\lambda_{A,B}(t,\tau)$ have been defined as a piecewise constant function so that the survival rate $P(X_t = A)$ exactly matches the data at the given points, with exponential decay in between.
  - Arbitrary data sources can be reproduced - care must be taken to avoid overfitting, typically by choosing appropriate parametric curves.

#### C.3 More complex hazards and infectious disease modelling

We can expand the types of models that can be simulated by allowing the hazard to depend on the current model state, i.e. have the form

$$\lambda_{i,j}(t,\tau,\mathbf{f}) \tag{35}$$

where $\mathbf{f}$ is the vector of density functions $f_i(t,\tau)$. This addition does not affect the results of Theorem 2.1, but does mean the method of characteristics describe in Section 2.2 many no longer be applied. The numerical scheme in Section 2.3 also needs adjustments; the scheme in equation (27) would not be valid in general, but (28) can be used to develop a new scheme, requiring estimation of the cumulative hazard integral in most cases.

By including this dependence in the hazard, one can simulate models where the effect size is dependent on the current state of the system. For example, in infection models, the risk of infection depends on the proportion of the population currently infected. We can write a standard SIR model[24] with states $S$ (susceptible), $I$ (infectious), $R$ (recovered) in this form. The risk of infection is proportional to the current infected population, leading to the hazard

$$\begin{aligned} \lambda_{S,I}(t,\tau,\mathbf{f}) &= \beta P(X_t = I) \\ &= \beta \int_0^\infty f_I(t,\tau)d\tau \\ &= \beta g_I(t). \end{aligned} \tag{36}$$

In the standard SIR model, risk recovery for infected individuals is constant, i.e.

$$\lambda_{I,R}(t,\tau,\mathbf{f}) = \gamma. \tag{37}$$

One can then confirm using equation (11) that

$$\frac{dg_S}{dt} = -\beta g_S(t) g_I(t), \quad \frac{dg_I}{dt} = \beta g_S(t) g_I(t) - \gamma g_I(t), \quad \frac{dg_R}{dt} = \gamma g_I(t), \tag{38}$$

which is the standard SIR model. From here one could modify this to introduce more complex risks of infection and recovery - for instance, a distribution for time to disease recovery could be simulated by modifying $\lambda_{I,R}$ to be a function of $\tau$. This is illustrated in Figure C.10. At left is a standard SIR model; right is an SIR model which assumes no

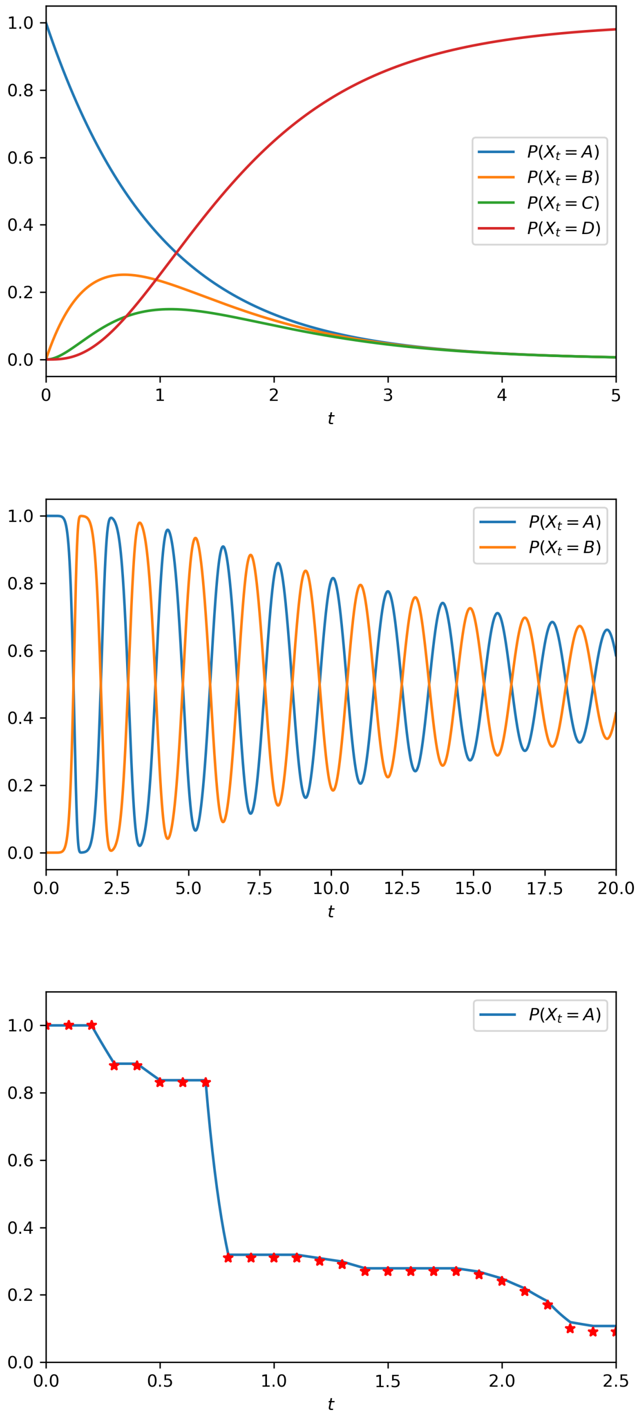


*Figure C.9: Example sojourn time density models, with differing dynamics.*

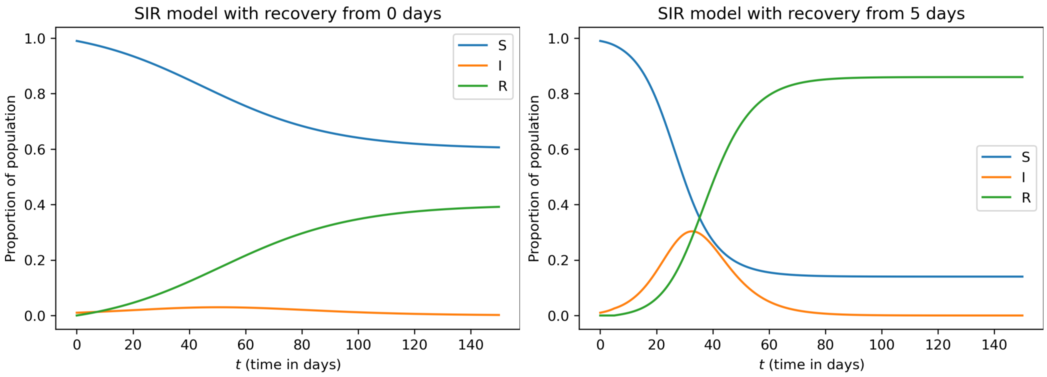


*Figure C.10: Sojourn time density models of a standard SIR simulation. Left - no delay before recovery. Right - 5 day delay before recovery.*

likelihood of recovery in the first five days of infection (i.e. $\lambda_{I,R}(t, \tau, \mathbf{f}) = 0$ if $\tau < 5$, $\gamma$ if $\tau \geq 5$). For this figure, $\beta = 0.2$, $\gamma = 0.16$. One can observe that if individuals cannot recover from infection, the total proportion of the population to ever be infected is much higher. This demonstrates the utility of the sojourn time density framework for this sort of system; new dynamics can be simulated by modifying the hazard functions, rather than the design of the model itself. One could further iterate, introducing an infection risk that varies with $t$, for instance.

The addition of heterogeneous generalises the sojourn time density model framework to incorporate many other types of model, including infectious disease models as illustrated above. This could allow for the synthesis of different model types into one framework; for example, a future model of liver disease which incorporates liver risk by hepatitis infection could include infectious disease modelling alongside the existing model of HCC and liver disease described in this manuscript.

#### C.4 Transitions in place

One of the major limitations of the sojourn time density models, in common with Markov and semi-Markov models, is that the sequence of transitions is memoryless, even if the time between transitions is not. There are a number of ways this has been addressed traditionally; for example, the introduction of hidden and augmented states.[25, 26, 27]. One way to address this in some circumstances is to add a transition which does not reset the sojourn time. For instance, in a model with three states $A_1, A_2, B$, where a transition from $A_1 \to A_2$ is independent of the transition to $B$, we may wish for the transition to not affect the likelihood of transition to $B$ and therefore to "conserve" the sojourn time in some sense.

To achieve this, we can introduce a new set of hazard rates $\mu_{i,j}(t, \tau)$ which are the transition rates for an individual in state $i$ to transition to state $j$, at time $t$, given that the individual has already spent time $\tau$ in the state without any transitions occurring, *while conserving the sojourn time*. Then equation (6) can be modified as follows:

$$\begin{aligned}\frac{\partial}{\partial t} f_i(t,\tau) + \frac{\partial}{\partial \tau} f_{i,j}(t,\tau) = &-\left(\sum_j \lambda_{i,j}(t,\tau)\right) f_i(t,\tau) \\ &-\left(\sum_j \mu_{i,j}(t,\tau)\right) f_i(t,\tau) + \left(\sum_k \mu_{k,i}(t,\tau) f_k(t,\tau)\right).\end{aligned} \tag{39}$$

Then Theorem 2.1 holds with minor alterations.

This approach may be useful for creating models equivalent to discrete event simulations with an "event queue" - if multiple events are queued at a given time, but a particular event does not reset the queue, this is equivalent to the sojourn time for that state "continuing"'.

### Appendix D CHEERS checklist

See Husereau et al for further details.[28]

| Topic | No. | Item | Location |
|---|---|---|---|
| Title | 1 | Identify the study as an economic evaluation and specify the interventions being compared. | Title |
| Abstract | 2 | Provide a structured summary that highlights context, key methods, results, and alternative analyses. | Abstract |
| **Introduction** | | | |
| Background and objectives | 3 | Give the context for the study, the study question, and its practical relevance for decision making in policy or practice. | Section 1 |
| **Methods** | | | |
| Health economic analysis plan | 4 | Indicate whether a health economic analysis plan was developed and where available. | N/A |
| Study population | 5 | Describe characteristics of the study population (such as age range, demographics, socioeconomic, or clinical characteristics). | Section 3.1, 3.7 |
| Setting and location | 6 | Provide relevant contextual information that may influence findings. | Section 3.1 |
| Comparators | 7 | Describe the interventions or strategies being compared and why chosen. | Section 3.4, 3.7 |
| Perspective | 8 | State the perspective(s) adopted by the study and why chosen. | Section 3.5 |
| Time horizon | 9 | State the time horizon for the study and why appropriate. | Section 3.5 |
| Discount rate | 10 | Report the discount rate(s) and reason chosen. | Section 3.5 |
| Selection of outcomes | 11 | Describe what outcomes were used as the measure(s) of benefit(s) and harm(s). | Section 3.5 |
| Measurement of outcomes | 12 | Describe how outcomes used to capture benefit(s) and harm(s) were measured. | Model outcome |
| Valuation of outcomes | 13 | Describe the population and methods used to measure and value outcomes. | Model outcome |
| Measurement, valuation of resources & costs | 14 | Describe how costs were valued. | Section 3.5 |
| Currency, price date, and conversion | 15 | Report the dates of the estimated resource quantities and unit costs, plus the currency and year of conversion. | Section 3.5, Table A.5 |
| Rationale and description of model | 16 | If modelling is used, describe in detail and why used. Report if the model is publicly available and where it can be accessed. | Section 2, 3. |
| Analytics and assumptions | 17 | Describe any methods for analysing or statistically transforming data, any extrapolation methods, and approaches for validating any model used. | Sections 3, 4.1, Appendix A. |

Table continued on next page.

| **Topic** | **No.** | **Item** | **Location** |
|---|---|---|---|
| Characterising heterogeneity | 18 | Describe any methods used for estimating how the results of the study vary for subgroups. | N/A |
| Characterising distributional effects | 19 | Describe how impacts are distributed across different individuals or adjustments made to reflect priority populations. | Only priority population modelled. |
| Characterising uncertainty | 20 | Describe methods to characterise any sources of uncertainty in the analysis. | Appendix B. |
| Approach to engagement with patients and others affected by the study | 21 | Describe any approaches to engage patients or service recipients, the general public, communities, or stakeholders (such as clinicians or payers) in the design of the study. | Section 1.4 |
| **Results** | | | |
| Study parameters | 22 | Report all analytic inputs (such as values, ranges, references) including uncertainty or distributional assumptions. | Appendix A |
| Summary of main results | 23 | Report the mean values for the main categories of costs and outcomes of interest and summarise them in the most appropriate overall measure. | Section 4.2, Table 1 |
| Effect of uncertainty | 24 | Describe how uncertainty about analytic judgments, inputs, or projections affect findings. Report the effect of choice of discount rate and time horizon, if applicable. | Appendix B |
| Effect of engagement with patients and others affected by the study | 25 | Report on any difference patient/service recipient, general public, community, or stakeholder involvement made to the approach or findings of the study | N/A |
| **Discussion** | | | |
| Study findings, limitations, generalisability, current knowledge | 26 | Report key findings, limitations, ethical or equity considerations not captured, and how these could affect patients, policy, or practice. | Section 5, Section 6 |
| **Other relevant information** | | | |
| Source of funding | 27 | Describe how the study was funded and any role of the funder in the identification, design, conduct, and reporting of the analysis | Funding information |
| Conflicts of interest | 28 | Report authors conflicts of interest according to journal or International Committee of Medical Journal Editors requirements. | Conflicts of interest |
